# Amygdala and Subgenual Cingulate Stressor-Evoked Activity Varies Across a Spectrum of Childhood Adversity Severity: Links to Affective and Cardiovascular Outcomes

**DOI:** 10.64898/2026.08.20.26360904

**Authors:** Nithya P. Kasibhatla, Christine W. Peng, Helmet T. Karim, Anusha Rangarajan, Nicholas A. Harris, Brandon M. Sibbach, Meredith L. Wallace, Howard J. Aizenstein, Layla Banihashemi

**Author notes:** Correspondence to: Layla Banihashemi, Ph.D. University of Pittsburgh Department of Psychiatry 3811 O’Hara St Pittsburgh, PA 15213.

## Abstract

**Background:** Childhood adversity is linked to psychopathology risk and dysregulated stress reactivity; however, unified underlying neural mechanisms are unclear. A “central visceral” network, including the bed nucleus of the stria terminalis (BNST), amygdala and subgenual anterior cingulate cortex (sgACC), is implicated in affective processes and proximally controls stress reactivity. We examined relationships among childhood adversity, stressor-evoked neural activity/connectivity and affective and cardiovascular outcomes.

**Methods:** Participants were adults (n=97, mean age=27.32, SD=4.02, 57 females) uniformly distributed across physical abuse severity. Childhood adversity was assessed by threat (abuse or traumatic events) and socioeconomic deprivation (SED). Participants performed an fMRI stress task with cardiovascular recordings.

Linear/curvilinear regressions were performed with threat and deprivation together as predictors of stressor-evoked activity/connectivity. Neural variables showing significant adversity associations were examined as predictors of affective symptoms/diagnoses or cardiovascular responses.

**Results:** Threat and SED displayed opposing curvilinear relationships with stressor-evoked amygdala and sgACC activity, respectively: at low and high adversity, amygdala reactivity was greater, whereas sgACC reactivity was blunted. Greater SED was associated with weaker BNST-sgACC connectivity. Blunted amygdala reactivity and lower sgACC reactivity were associated with greater post-traumatic stress symptoms. Affective diagnoses peaked at near-zero BNST-sgACC connectivity. Greater amygdala reactivity was associated with blunted diastolic blood pressure reactivity and recovery.

**Conclusions:** The curvilinear relationships suggest adversity-related vulnerability thresholds. Blunted amygdala, lower sgACC reactivity and weaker BNST-sgACC connectivity may confer affective risk, whereas heightened amygdala reactivity may confer cardiovascular risk. Our findings support a central visceral network pathway by which childhood adversity may contribute to affective and cardiovascular health.

## Introduction

Childhood adversity is a dimensional construct comprising threat (events presenting hazards to physical integrity) and deprivation (the absence of expected environmental stimuli/exposures)(1, 2); a large literature demonstrates brain structural and functional consequences of adversity(3–7). Reports indicate high global prevalence of violence toward children(8) and aggressive physical and psychological discipline(9). Childhood adversity is prospectively linked to adult mental health conditions, including mood, anxiety and trauma-related disorders(10–14), and to increased risk for physical health conditions, including cardiovascular disease(15–17). Understanding neural mechanisms underlying childhood adversity-related mental and physical health risks can improve preventative and therapeutic measures.

Childhood adversity may contribute to stress-related health problems by dysregulating physiological stress responses(18, 19) and is associated with dysregulated (heightened or blunted) neuroendocrine(20–23) and autonomic reactivity, including cardiovascular (heart rate (HR) and blood pressure) reactivity(21, 24–29). These relationships may vary by adversity dimension: childhood threat (i.e., abuse/maltreatment) is associated with blunted physiological stress reactivity(30–34), whereas deprivation (i.e., low socioeconomic status (SES)) is associated with heightened levels of basal stress markers and reactivity(35–38). This divergence is consistent with models of context-dependent, adversity-related calibration of stress reactivity(39). Despite mounting evidence linking childhood threat and deprivation to altered physiological stress systems, how childhood adversity impacts specific, proximal stress-regulatory neural circuits remains unclear.

Physiological stress responses are most proximally regulated by a core central visceral circuit (CVC), which receives and integrates viscerosensory input and controls autonomic outflow to viscera(40). CVCs include the paraventricular hypothalamus (PVN), bed nucleus of the stria terminalis (BNST), amygdala and subgenual anterior cingulate cortex (sgACC). The PVN, as the apex of the hypothalamic pituitary adrenal (HPA) axis, is the primary controller of neuroendocrine stress responses, and is poised to control autonomic outflow (sympathetic and parasympathetic) outflow(41, 42), including cardiovascular responses(43, 44). The PVN receives dense input from the BNST(45), with a more limited input from the central nucleus of the amygdala(46, 47), structures that are also directly interconnected(48–50). Reciprocal connections exist between sgACC and the extended amygdala, with direct projections from sgACC to BNST(51–53) and interconnections between sgACC and amygdala(54, 55).

CVCs are involved in affective processes, such as emotional memory, threat and fear responses and emotional regulation(56–61), and are implicated in psychopathology (e.g., depression, anxiety, trauma-related disorders)(62–71). These regions also exert direct control over cardiovascular function through neuroendocrine and preautonomic pathways(44, 72–74). They each innervate brainstem cardiorespiratory centers(75–77), where stress signaling and cardiovascular regulation converge.

Preclinical studies demonstrate that early experience shapes the structure and function of CVCs, influencing the neonatal synaptic assembly of preautonomic circuits(78), as well as preautonomic circuit strength(79) and stress-induced PVN and BNST activation later in life(80). Several human imaging studies have linked childhood adversity and stressor-evoked neural activity in central visceral regions using threat or stress tasks. One study found that maltreatment was associated with greater amygdala, but not BNST, activation during anticipated threat of shock(81). Another study linked greater maltreatment to greater psychosocial stressor-evoked insula and prefrontal activation(82). In a military veteran sample, childhood maltreatment moderated the association between combat exposure and stressor-evoked sgACC activity, among other cingulate subregions(83).

Relatively few human imaging studies have examined relationships between childhood adversity and stressor-evoked activity within this stress-proximal CVC network. One study examined CVC stressor-evoked activity in a healthy adult sample using interference-evoking MRI tasks commonly used to elicit subjective and physiological stress states(38, 84, 85); childhood threat (physical abuse) was significantly associated with stressor-evoked activity within the preautonomic hypothalamus/PVN, BNST, amygdala and sgACC(86). These results suggest that CVC regions may be sensitive to levels of abuse that fall below the severe-to-extreme range, characteristic of a healthy sample. Stressor-evoked activity within these CVC regions was also associated with HR reactivity, and our prior work has linked CVC regions to affective symptoms/disorders, including BNST-PVN resting-state connectivity and the visceral white matter of the medial forebrain bundle(87, 88).

Despite these findings indicating CVCs as a potential link between childhood adversity and cardiovascular and affective outcomes, no studies have investigated CVC stressor-evoked activity/connectivity as an underlying neural mechanism in an abuse-enriched, transdiagnostic sample. Further, of the studies examining childhood adversity and neural stressor-evoked activity, few have examined threat and deprivation simultaneously. Moreover, prior work has largely focused on linear relationships, potentially obscuring non-linear patterns wherein moderate versus extreme adversity may differentially affect CVC function and downstream affective and cardiovascular outcomes. Recent neuroimaging work has demonstrated curvilinear associations between adversity and brain function(89, 90), and between threat-evoked brain activity and peripheral physiological responses(91, 92).

The current study expands previous work with a transdiagnostic, abuse-enriched sample to further elucidate linear and curvilinear relationships between 1) childhood adversity and CVC stressor-evoked activity and connectivity, and between these CVC measures and both 2) affective symptoms and diagnoses and 3) cardiovascular stress responses. Building on our prior dimensional findings (opposing relationships of childhood threat and deprivation with visceral white matter structural integrity, and threat-specific relationships with CVC resting-state connectivity(87, 88)) and extending this framework to stressor-evoked neural activity, we hypothesized that childhood threat would be associated with blunted, while childhood deprivation would be associated with heightened, CVC stressor-evoked activity. We further hypothesized that adversity-related differences in CVCs would, in turn, be associated with affective symptom severity and cardiovascular stress reactivity. Our hypothesis reflects a candidate neural mechanism whereby childhood adversity may affect mental and physical health through upstream dysregulation of CVCs. Elucidating how CVCs may contribute to both physiological and affective outcomes may aid translation from preclinical research and provide novel, proximally stress-responsive targets and interventions for adversity-related health conditions.

## Methods and Materials

### Participants

Participants were recruited from Allegheny County, Pennsylvania via research study referrals and bus/online advertisements. Of 111 consented, 100 participants completed study procedures. Participants were 59 female and 41 male young adults (n=100, mean age=27.28, SD=3.99); self-reported race was 13% Asian, 2% biracial, 36% Black or African American, 4% multiracial and 45% White. The University of Pittsburgh Institutional Review Board approved study protocols and all participants provided informed consent.

Standard MRI, psychiatric and medical exclusion criteria were applied as previously described(87, 88) (Supplement). Participants with depression, anxiety or trauma-related disorders were eligible.

Recruitment targeted a relatively even distribution of physical abuse severity, using the Childhood Trauma Questionnaire (CTQ) physical abuse subscale as a screening measure(93). The final sample (n=100) reflected this: 29% None-Minimal, 23% Low-Moderate, 21% Moderate-Severe, and 27% Severe-Extreme.

### Study Protocol and Measures

The study comprised two visits (mean interval: 14.39 ± 10.96 days), an intake visit followed by an MRI scan visit at the University of Pittsburgh Magnetic Resonance Research Center.

### Childhood Threat

Childhood threat was assessed with the CTQ and the Trauma History Questionnaire (THQ). The CTQ examines physical, emotional and sexual abuse, and physical and emotional neglect(93). CTQ Threat is the sum of the three abuse subscales (with 15 indicating no abuse and 75 indicating extreme abuse). (CTQ Deprivation is the sum of the two neglect subscales.)

The THQ assesses the occurrence of traumatic events throughout the life course(94); an adapted version indicates occurrence, repetition, and age range(s): age 0-11, 12-17, and >18(95). THQ 0-11 indexed childhood threat; adulthood traumatic events (THQ>18) served as a covariate.

### Childhood Socioeconomic Deprivation

Maximum parental education level and the participants’ education level were indices of childhood and adulthood SES, respectively. Both were rated on a 9-point scale (0–No high school diploma, 8–Doctorate). SES and socioeconomic deprivation (SED) are included in the Childhood Deprivation construct(2, 96–98) and education is a common SES measure(99–101). As in prior work(87, 88), given the strong correlation between CTQ Threat and CTQ Deprivation (Supplement, Table S1), we used parental education level (reverse coded) as our measure of deprivation. Adulthood education served as a SES variable/covariate.

### Negative Life Events

The Life Events List captured major life events occurring in the past year (e.g., unemployment, divorce or separation, serious illness, death of a loved one)(102); we used the resulting count of negative life events as a covariate.

### Sample Characterization

Sample characterization measures included the Perceived Stress Scale (PSS, 10-item version)(103), the State Trait Anxiety Inventory (STAI-Y2)(104) and the 60-item NEO Five-Factor Inventory-3 (NEO-FFI-3)(105) (Table S2).

### fMRI Stressor Task: Multisource Interference Task

During the Multisource Interference Task (MSIT), participants identify the number that differs among three numbers via corresponding glove positions. In the congruent (control) condition, the target’s location matches its response position; in the incongruent condition, the two are mismatched, with accuracy titrated to ∼50% by adjusting the inter-trial interval. Task parameters were as previously described(86, 106–108) (Supplement). The MSIT resulted in significant anxious arousal, less perceived control and less positive valence (Supplement, Fig. S1).

### MRI Protocol and Data Acquisition

MRI data were collected on a 3-Tesla Trio TIM MRI scanner (Siemens, Erlangen, Germany) with a 32-channel head coil. See Supplement for acquisition parameters. The MSIT sequence followed MPRAGE and resting-state acquisitions and preceded diffusion spectrum imaging, reported previously(87, 88) (total scan ∼50–55 minutes).

### Preprocessing

fMRI data were preprocessed and analyzed using Statistical Parametric Mapping software (SPM12, http://www.fil.ion.ucl.ac.uk/spm/). (See Supplement for details.)

### Level 1 Analyses

Contrast maps reflecting relative task-related blood oxygen level–dependent (BOLD) signal change between congruent and incongruent conditions were estimated per participant using first-level GLM analysis (Supplement). Data from 97 participants were analyzed; three were excluded for excessive movement (>3mm, any axis/rotation).

### Level 2 Analysis

To examine main effects of the MSIT, a whole-brain Level 2 analysis was conducted (Supplement, Fig. S2).

### Region of Interest Analyses

Relative signal change within the four regions of interest (ROIs)(86) was estimated per task condition using small volume correction (whole-brain maps masked by each ROI, FDR<0.05), extracting the first eigenvariate from each significant cluster (Supplement). Significant deactivation (congruent>incongruent) was detected in all four ROIs; significant activation (incongruent>congruent) was detected only in the amygdala. Consistent with prior work showing stressor-evoked *deactivation* within these ROIs(86), analyses used only deactivation clusters, maintaining a single interpretive convention (greater deactivation indicating greater reactivity; Supplement, Fig. S3).

### Stressor-Evoked Connectivity: Generalized Psychophysiological Interaction Analyses (gPPI)

Task-dependent connectivity was estimated using the SPM8 Generalized PPI Toolbox(109) from each ROI seed to the rest of the brain; each seed’s activation map was then masked with each other ROI to generate significant PPI clusters. *Negative* connectivity values indicate *greater* task-related coupling during incongruent compared to congruent conditions; *positive* values indicate *greater* coupling during congruent compared to incongruent conditions; near-zero connectivity reflects minimal differential coupling between conditions.

Significant connectivity in the congruent condition was found between all ROI pairs (Supplement).

### Affective Measures

#### Affective Symptom Severity

Depression and post-traumatic stress symptom (PTSS) severity were assessed using the Beck Depression Inventory (BDI-II)(110) and the post-traumatic stress disorder (PTSD) Checklist - Civilian Version (PCL-C)(111), respectively.

#### Diagnostic Assessment

Psychiatric diagnoses of mood, anxiety or trauma-related disorders were evaluated via in-person interview using the Structured Clinical Interview for DSM-IV Axis I Disorders (LB). Of the 97 participants, 28 were healthy and 69 had a history of affective diagnosis (Supplement).

### Cardiovascular

#### Cardiovascular Measures

A MagLife Light MRI Patient Monitor recorded systolic (SBP) and diastolic blood pressure (DBP) and HR via an oscillometric cuff placed on the participant’s left arm. Recordings were at one-minute intervals during the MPRAGE and MSIT sequences and two-minute intervals during the final sequence (diffusion spectrum imaging, 19-min) for baseline, stress and post-stress recovery measures, respectively.

#### Cardiovascular Outcomes

Cardiovascular measures (SBP, DBP and HR) were averaged across 4 baseline, 4 incongruent (stress), and 6 post-stress recovery recordings. Stress reactivity was calculated as averaged stress minus averaged baseline; recovery as averaged recovery minus averaged stress.

### Data Analysis

#### Childhood Adversity and Affective or Cardiovascular Outcomes

Bivariate Pearson correlations examined linear relationships between childhood adversity variables and affective and cardiovascular outcomes.

#### Childhood Adversity and CVC Stressor-Evoked Activity/Connectivity

We examined additive effects of childhood threat and SED(112) on stressor-evoked neural measures (activity and connectivity) using hierarchical regression. Because childhood threat measures were highly correlated (r=0.618, Table S1), CTQ Threat and THQ 0-11 were examined in separate models, as previously(87, 88).

Each hierarchical model included: Step 1: age, race and sex; Step 2: CTQ Threat or THQ 0-11, with SED; Step 3: adulthood covariates (THQ >18, adulthood education, and negative life events). Multiple comparison correction was conducted (FDR <0.05; four tests for stressor-evoked activity, seven tests for connectivity)(113). Curvilinear models included squared adversity terms.

#### CVC Stressor-Evoked Activity/Connectivity and Affective or Cardiovascular Outcomes

To identify potential CVC links between childhood adversity and outcomes, we examined whether adversity-associated neural measures predicted affective symptoms (BDI-II, PCL-C, or the number of lifetime diagnoses) or cardiovascular outcomes (SBP, DBP and HR baseline, stress reactivity and recovery). We used the same hierarchical regression approach described above, with stressor-evoked neural measures as predictors and affective or cardiovascular outcomes as dependent variables. Multiple comparison correction was applied for affective outcomes (FDR <0.05, three tests) and each cardiovascular parameter (FDR <0.05, three tests, separately for DBP, SBP and HR). Standardized β values are reported as effect size; for curvilinear effects, β reflects the quadratic term.

## Results

### Childhood Adversity and Affective or Cardiovascular Outcomes

See Supplement for correlations between childhood adversity and affective (Table S3) and cardiovascular (Table S4) outcomes.

### Childhood Threat, Deprivation and CVC Stressor-Evoked Activity

#### Abuse (CTQ Threat)

In the abuse models, analyses revealed a significant inverted U-shaped curvilinear relationship between CTQ Threat and stressor-evoked amygdala activity (β=-1.352, *P=*0.012, Table 1, Figure 1A) (no significant effect of SED). This relationship remained significant in the full model with the adulthood variables (THQ>18, adulthood SES and negative life events) (β=-1.627, *P=*0.003). Analyses also revealed a significant U-shaped relationship between SED and stressor-evoked sgACC activity (β=1.199, *P=*0.018, Figure 1B) (no significant effect of CTQ Threat). This relationship remained significant in the full model (β=1.121, *P=*0.031, Table 1).

**Figure 1.**
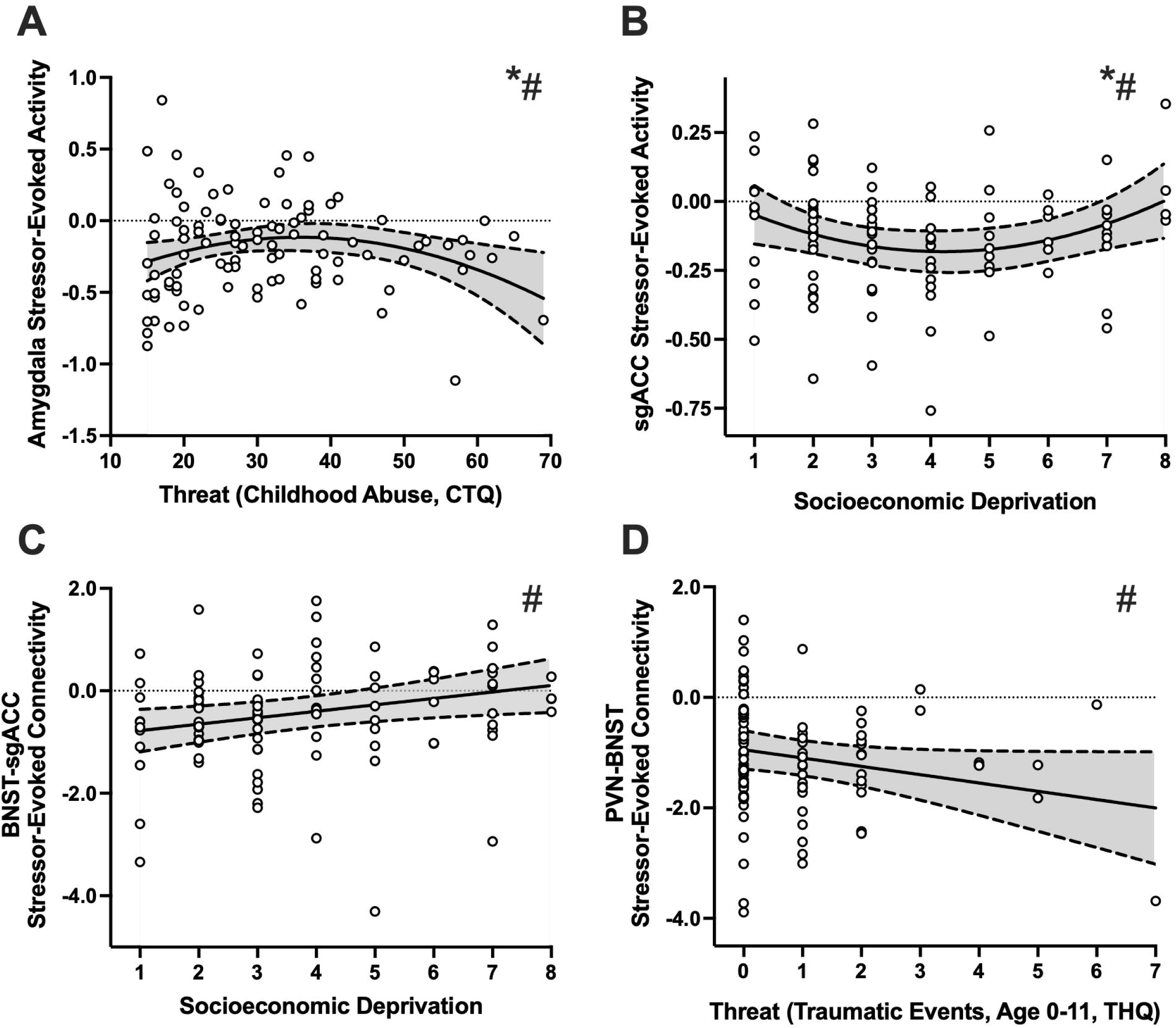
Childhood threat and deprivation are associated with Central Visceral Circuit stressor-evoked activity and connectivity. Hierarchical linear regression was used to examine the additive effects of childhood threat and socioeconomic deprivation (SED) on CVC stressor-evoked activity and connectivity (Control>Incongruent contrast), tested in separate models using childhood abuse (Childhood Trauma Questionnaire, CTQ Threat) and early, repeated traumatic events (Trauma History Questionnaire, THQ 0–11) as threat variables. **(A)** CTQ Threat showed a significant inverted U-shaped relationship with amygdala activity, with low and high threat associated with greater deactivation (reactivity) and moderate threat associated with more blunted (near-zero) reactivity (β=-1.352, *P=*0.012). **(B)** SED showed a significant U-shaped relationship with sgACC activity, with low and high deprivation associated with more blunted reactivity and moderate deprivation associated with greater reactivity (CTQ Threat model: β=1.199, *P=*0.018; THQ 0-11 model: β=1.237, *P=*0.011). **(C)** SED was linearly associated with BNST-sgACC connectivity, such that greater SED was associated with weaker (near-zero) connectivity (CTQ Threat model: β=0.301, *P=*0.008, THQ 0-11 model: β=0.262, *P=*0.016). **(D)** THQ 0–11 was linearly associated with PVN-BNST connectivity, such that greater trauma was associated with greater connectivity (β=-0.226, *P=*0.048). *Indicates survival of FDR correction (0.05, four tests for stressor-evoked activity, seven tests for each region of interest to region of interest connection); (B) survived FDR correction within the THQ 0–11 model. #Indicates the association was significant in the full model with the adulthood covariates (traumatic events >18 years, education level/SES, past-year negative life events). (BNST, bed nucleus of the stria terminalis; PVN, paraventricular nucleus of the hypothalamus; sgACC, subgenual anterior cingulate cortex).

**Table 1.** Regression Results: CTQ Threat (Abuse) and Stressor-Evoked Activity.

| Step | Variable | Amygdala |  |  | sgACC |  |  | PVN |  |  | BNST |  |  |
| --- | --- | --- | --- | --- | --- | --- | --- | --- | --- | --- | --- | --- | --- |
| | | St. $\beta$ | t | p | St. $\beta$ | t | p | St. $\beta$ | t | p | St. $\beta$ | t | p |
| 1 | Age | -.044 | -.425 | .672 | .060 | .571 | .569 | .050 | .478 | .634 | .000 | -.003 | .998 |
|  | Sex | .123 | 1.193 | .236 | .030 | .290 | .773 | .079 | .764 | .447 | .114 | 1.103 | .273 |
|  | Race | .127 | 1.238 | .219 | .071 | .681 | .497 | -.052 | -.502 | .617 | -.043 | -.419 | .676 |
| 2 | Age | -.052 | -.480 | .632 | .045 | .403 | .688 | .032 | .286 | .776 | -.018 | -.161 | .872 |
|  | Sex | .123 | 1.162 | .248 | .040 | .368 | .713 | .087 | .817 | .416 | .120 | 1.126 | .263 |
|  | Race | .139 | 1.323 | .189 | .069 | .642 | .522 | -.045 | -.428 | .670 | -.032 | -.301 | .764 |
|  | CTQ Threat | -.052 | -.445 | .657 | .029 | .245 | .807 | -.011 | -.093 | .926 | -.036 | -.310 | .757 |
|  | Socioeconomic Deprivation | .079 | .688 | .493 | .030 | .254 | .800 | .078 | .669 | .505 | .098 | .845 | .400 |
| 3 | Age | -.044 | -.410 | .683 | .090 | .821 | .414 | .026 | .232 | .817 | -.037 | -.326 | .745 |
|  | Sex | .122 | 1.181 | .241 | .045 | .432 | .667 | .086 | .797 | .428 | .117 | 1.093 | .277 |
|  | Race | .095 | .919 | .360 | .054 | .508 | .613 | -.054 | -.494 | .623 | -.033 | -.309 | .758 |
|  | CTQ Threat | 1.298 | 2.443 | **.017** | .080 | .147 | .883 | .304 | .545 | .587 | .190 | .343 | .732 |
|  | Socioeconomic Deprivation | -.654 | -1.318 | .191 | -1.165 | -2.305 | **.024** | .093 | .179 | .858 | .498 | .963 | .338 |
|  | **CTQ Threat<sup>2</sup>** | -1.352 | -2.578 | **.012\*** | -.018 | -.034 | .973 | -.320 | -.582 | .562 | -.241 | -.441 | .660 |
|  | **Socioeconomic Deprivation<sup>2</sup>** | .745 | 1.534 | .129 | 1.199 | 2.421 | **.018** | -.013 | -.026 | .979 | -.400 | -.789 | .432 |
| 4 | Age | .067 | .561 | .576 | .139 | 1.117 | .267 | .105 | .824 | .412 | .062 | .496 | .621 |
|  | Sex | .098 | .913 | .364 | .039 | .353 | .725 | .061 | .536 | .594 | .110 | .973 | .333 |
|  | Race | .094 | .898 | .372 | .039 | .363 | .718 | -.056 | -.507 | .614 | -.025 | -.225 | .822 |
|  | CTQ Threat | 1.620 | 2.925 | **.004** | .205 | .358 | .721 | .543 | .923 | .358 | .461 | .795 | .429 |
|  | Socioeconomic Deprivation | -.610 | -1.227 | .223 | -1.115 | -2.165 | **.033** | .138 | .261 | .795 | .494 | .949 | .345 |
|  | **CTQ Threat<sup>2</sup>** | -1.627 | -3.010 | **.003\*** | -.121 | -.217 | .829 | -.524 | -.913 | .364 | -.475 | -.840 | .403 |
|  | **Socioeconomic Deprivation<sup>2</sup>** | .728 | 1.478 | .143 | 1.121 | 2.199 | **.031** | -.038 | -.072 | .942 | -.356 | -.691 | .492 |
|  | THQ >18 | -.238 | -1.830 | .071 | -.099 | -.731 | .467 | -.154 | -1.111 | .270 | -.242 | -1.774 | .080 |
|  | Adulthood SES | -.056 | -.502 | .617 | -.113 | -.980 | .330 | -.043 | -.364 | .717 | .002 | .018 | .985 |
|  | Negative Life Events | -.044 | -.365 | .716 | -.014 | -.113 | .910 | -.063 | -.492 | .624 | .017 | .133 | .895 |

Bold values indicate significance at $p < 0.05$ ; an asterisk indicates survival of FDR correction (0.05) for 4 tests. BNST, bed nucleus of the stria terminalis; PVN, paraventricular nucleus of the hypothalamus; sgACC, subgenual anterior cingulate cortex; CTQ, Childhood Trauma Questionnaire; SES, Socioeconomic Status; THQ, Trauma History Questionnaire.

#### Early, Repeated Traumatic Events (THQ 0-11)

Similar results were observed in the THQ 0-11 models; THQ 0-11 showed a significant inverted U-shaped relationship with amygdala activity (β=-0.546, *P=*0.033, Figure S4A). (Potential outliers are reported in the Supplement, Table S5.) SED showed a significant U-shaped relationship with sgACC activity (β=1.237, *P=*0.011, Table 2), both remaining significant in the full model.

**Table 2.** Regression Results: Early, Repeated Traumatic Events (age 0-11) and Stressor-Evoked Activity.

| Step | Variable | Amygdala |  |  | sgACC |  |  | PVN |  |  | BNST |  |  |
| --- | --- | --- | --- | --- | --- | --- | --- | --- | --- | --- | --- | --- | --- |
| | | St. $\beta$ | t | p | St. $\beta$ | t | p | St. $\beta$ | t | p | St. $\beta$ | t | p |
| 1 | Age | -.044 | -.425 | .672 | .060 | .571 | .569 | .050 | .478 | .634 | .000 | -.003 | .998 |
|  | Sex | .123 | 1.193 | .236 | .030 | .290 | .773 | .079 | .764 | .447 | .114 | 1.103 | .273 |
|  | Race | .127 | 1.238 | .219 | .071 | .681 | .497 | -.052 | -.502 | .617 | -.043 | -.419 | .676 |
| 2 | Age | -.058 | -.538 | .592 | .048 | .444 | .658 | .034 | .318 | .751 | -.020 | -.189 | .851 |
|  | Sex | .131 | 1.260 | .211 | .035 | .332 | .741 | .090 | .865 | .389 | .126 | 1.205 | .231 |
|  | Race | .144 | 1.384 | .170 | .071 | .666 | .507 | -.024 | -.229 | .819 | -.022 | -.213 | .832 |
|  | THQ 0-11 | -.100 | -.950 | .345 | .020 | .188 | .852 | -.177 | -1.680 | .096 | -.117 | -1.109 | .270 |
|  | Socioeconomic Deprivation | .081 | .739 | .462 | .036 | .320 | .749 | .108 | .992 | .324 | .108 | .985 | .327 |
| 3 | Age | -.036 | -.333 | .740 | .102 | .944 | .348 | .023 | .214 | .831 | -.041 | -.373 | .710 |
|  | Sex | .123 | 1.198 | .234 | .031 | .299 | .766 | .094 | .900 | .371 | .125 | 1.188 | .238 |
|  | Race | .131 | 1.278 | .205 | .052 | .505 | .615 | -.017 | -.167 | .868 | -.017 | -.163 | .871 |
|  | THQ 0-11 | .417 | 1.629 | .107 | .355 | 1.380 | .171 | -.426 | -1.627 | .107 | -.095 | -.361 | .719 |
|  | Socioeconomic Deprivation | -.399 | -.825 | .412 | -1.206 | -2.486 | **.015** | .338 | .685 | .495 | .606 | 1.219 | .226 |
|  | **THQ 0-11<sup>2</sup>** | -.546 | -2.165 | **.033** | -.325 | -1.286 | .202 | .263 | 1.020 | .310 | -.039 | -.150 | .881 |
|  | **Socioeconomic Deprivation<sup>2</sup>** | .459 | .968 | .336 | 1.237 | 2.599 | **.011\*** | -.220 | -.455 | .651 | -.503 | -1.033 | .305 |
| 4 | Age | .012 | .099 | .921 | .151 | 1.206 | .231 | .071 | .560 | .577 | .033 | .261 | .795 |
|  | Sex | .136 | 1.235 | .220 | .028 | .256 | .798 | .077 | .678 | .500 | .128 | 1.136 | .259 |
|  | Race | .135 | 1.280 | .204 | .041 | .385 | .701 | -.017 | -.161 | .872 | -.003 | -.032 | .974 |
|  | THQ 0-11 | .449 | 1.723 | .088 | .368 | 1.408 | .163 | -.423 | -1.582 | .117 | -.059 | -.222 | .825 |
|  | Socioeconomic Deprivation | -.402 | -.809 | .421 | -1.149 | -2.305 | **.024** | .392 | .769 | .444 | .610 | 1.199 | .234 |
|  | **THQ 0-11<sup>2</sup>** | -.562 | -2.135 | **.036** | -.306 | -1.161 | .249 | .298 | 1.104 | .273 | -.041 | -.153 | .879 |
|  | **Socioeconomic Deprivation<sup>2</sup>** | .462 | .941 | .350 | 1.151 | 2.339 | **.022** | -.263 | -.524 | .602 | -.477 | -.949 | .345 |
|  | THQ >18 | -.140 | -1.053 | .295 | -.102 | -.763 | .447 | -.085 | -.621 | .536 | -.194 | -1.426 | .157 |
|  | Adulthood SES | -.028 | -.242 | .809 | -.109 | -.950 | .345 | -.031 | -.263 | .793 | .015 | .131 | .896 |
|  | Negative Life Events | .076 | .586 | .560 | .008 | .065 | .949 | -.041 | -.304 | .762 | .058 | .435 | .664 |

Bold values indicate significance at $p < 0.05$ ; an asterisk indicates survival of FDR correction (0.05) for 4 tests. BNST, bed nucleus of the stria terminalis; PVN, paraventricular nucleus of the hypothalamus; sgACC, subgenual anterior cingulate cortex; SES, Socioeconomic Status; THQ, Trauma History Questionnaire.

### Childhood Threat, Deprivation and CVC Stressor-Evoked Connectivity

#### Abuse (CTQ Threat)

In the abuse models, analyses revealed significant linear effects of SED on stressor-evoked connectivity (Table 3): Amygdala-BNST (β=0.243, *P=*0.035), BNST-sgACC (β=0.301, *P=*0.008), PVN-BNST (β=0.264, *P=*0.021), sgACC-BNST (β=0.252, *P=*0.027), with no significant effects of abuse (Table 3). The relationship between SED and BNST-sgACC stressor-evoked connectivity remained significant in the full model (β=0.284, *P=*0.023, Figure 1C) while effects on PVN-BNST and sgACC-BNST became trends. The association between SED and sgACC-BNST stressor-evoked connectivity was significant in the curvilinear model (U-shaped, β=1.067, *P=*0.028, Figure S4B) and remained significant in the full model (β=1.068, *P=*0.033, data not shown).

**Table 3.** Regression Results: CTQ Threat (Abuse) and Stressor-Evoked Connectivity (Abbreviated)

| Step | Variable | Amygdala-BNST |  |  | BNST-sgACC |  |  | PVN-BNST |  |  | sgACC-BNST |  |  |
| --- | --- | --- | --- | --- | --- | --- | --- | --- | --- | --- | --- | --- | --- |
| | | St. $\beta$ | t | p | St. $\beta$ | t | p | St. $\beta$ | t | p | St. $\beta$ | t | p |
| 1 | Age | -.102 | -.986 | .326 | -.166 | -1.608 | .111 | -.123 | -1.191 | .237 | -.143 | -1.384 | .170 |
|  | Sex | -.069 | -.668 | .506 | .009 | .089 | .930 | -.061 | -.593 | .555 | -.090 | -.878 | .382 |
|  | Race | -.050 | -.482 | .631 | .070 | .681 | .498 | -.039 | -.373 | .710 | .008 | .076 | .939 |
| 2 | Age | -.160 | -1.484 | .141 | -.210 | -2.000 | .048 | -.153 | -1.435 | .155 | -.172 | -1.617 | .109 |
|  | Sex | -.044 | -.417 | .677 | .020 | .196 | .845 | -.059 | -.572 | .569 | -.088 | -.848 | .399 |
|  | Race | -.029 | -.284 | .777 | .110 | 1.087 | .280 | .002 | .023 | .982 | .046 | .452 | .653 |
|  | **CTQ Threat** | -.033 | -.286 | .776 | -.149 | -1.335 | .185 | -.169 | -1.497 | .138 | -.158 | -1.401 | .165 |
|  | **Socioeconomic Deprivation** | .243 | 2.146 | **.035** | .301 | 2.725 | **.008** | .264 | 2.351 | **.021** | .252 | 2.254 | **.027** |
| 3 | Age | -.147 | -1.195 | .235 | -.221 | -1.835 | .070 | -.151 | -1.243 | .217 | -.131 | -1.082 | .282 |
|  | Sex | -.022 | -.198 | .843 | .013 | .121 | .904 | -.022 | -.205 | .838 | -.085 | -.778 | .439 |
|  | Race | -.039 | -.365 | .716 | .101 | .965 | .337 | -.004 | -.041 | .967 | .039 | .368 | .714 |
|  | **CTQ Threat** | -.030 | -.256 | .798 | -.149 | -1.285 | .202 | -.176 | -1.503 | .137 | -.140 | -1.195 | .235 |
|  | **Socioeconomic Deprivation** | .206 | 1.651 | .102 | .284 | 2.318 | **.023** | .225 | 1.820 | .072 | .232 | 1.878 | .064 |
|  | THQ > 18 | -.056 | -.425 | .672 | .049 | .379 | .705 | -.062 | -.474 | .637 | -.089 | -.681 | .498 |
|  | Adulthood SES | -.078 | -.695 | .489 | -.037 | -.333 | .740 | -.063 | -.568 | .571 | -.078 | -.698 | .487 |
|  | Negative Life Events | .077 | .627 | .532 | -.037 | -.303 | .763 | .134 | 1.098 | .275 | .015 | .121 | .904 |

Bold values indicate significance at $p < 0.05$ ; findings did not survive FDR correction (0.05) for 7 tests. BNST, bed nucleus of the stria terminalis; PVN, paraventricular nucleus of the hypothalamus; sgACC, subgenual anterior cingulate cortex; CTQ, Childhood Trauma Questionnaire; SES, Socioeconomic Status; THQ, Trauma History Questionnaire.

#### Early, Repeated Traumatic Events (THQ 0-11)

Analyses revealed a linear relationship between THQ 0-11 and PVN-BNST that was significant in the full model (β=-0.226, *P=*0.048, Table 4, Figure 1D). Relationships between SED and stressor-evoked connectivity in the THQ 0-11 models were similar to the abuse models (Table 4), with only BNST-sgACC remaining significant in the full model (β=0.245, *P=*0.042). The U-shaped relationship between SED and sgACC-BNST stressor-evoked connectivity (described above) was also significant in the trauma model (β=1.071, *P=*0.022), remaining significant in the full model (β=1.077, *P=*0.027, data not shown).

**Table 4.** Regression Results: Early, Repeated Traumatic Events (age 0-11) and Stressor-Evoked Connectivity (Abbreviated)

| Step | Variable | Amygdala-BNST |  |  | BNST-sgACC |  |  | PVN-BNST |  |  | sgACC-BNST |  |  |
| --- | --- | --- | --- | --- | --- | --- | --- | --- | --- | --- | --- | --- | --- |
| | | St. $\beta$ | t | p | St. $\beta$ | t | p | St. $\beta$ | t | p | St. $\beta$ | t | p |
| 1 | Age | -.102 | -.986 | .326 | -.166 | -1.608 | .111 | -.123 | -1.191 | .237 | -.143 | -1.384 | .170 |
|  | Sex | -.069 | -.668 | .506 | .009 | .089 | .930 | -.061 | -.593 | .555 | -.090 | -.878 | .382 |
|  | Race | -.050 | -.482 | .631 | .070 | .681 | .498 | -.039 | -.373 | .710 | .008 | .076 | .939 |
| 2 | Age | -.162 | -1.535 | .128 | -.231 | -2.210 | .030 | -.174 | -1.662 | .100 | -.192 | -1.833 | .070 |
|  | Sex | -.038 | -.373 | .710 | .042 | .417 | .678 | -.033 | -.327 | .744 | -.063 | -.624 | .534 |
|  | Race | -.023 | -.225 | .822 | .094 | .928 | .356 | .000 | -.003 | .998 | .043 | .427 | .671 |
|  | **THQ 0-11** | -.088 | -.845 | .400 | -.062 | -.602 | .549 | -.188 | -1.829 | .071 | -.171 | -1.664 | .099 |
|  | **Socioeconomic Deprivation** | .248 | 2.310 | **.023** | .262 | 2.463 | **.016** | .242 | 2.272 | **.025** | .231 | 2.171 | **.033** |
| 3 | Age | -.166 | -1.343 | .183 | -.237 | -1.929 | .057 | -.198 | -1.631 | .107 | -.166 | -1.350 | .180 |
|  | Sex | -.008 | -.071 | .943 | .034 | .313 | .755 | .020 | .181 | .857 | -.054 | -.495 | .622 |
|  | Race | -.031 | -.297 | .767 | .085 | .808 | .421 | -.004 | -.043 | .966 | .036 | .347 | .730 |
|  | **THQ 0-11** | -.102 | -.894 | .374 | -.058 | -.509 | .612 | -.226 | -2.008 | **.048** | -.159 | -1.403 | .164 |
|  | **Socioeconomic Deprivation** | .209 | 1.747 | .084 | .245 | 2.060 | **.042** | .198 | 1.678 | .097 | .208 | 1.753 | .083 |
|  | THQ > 18 | -.032 | -.242 | .810 | .040 | .303 | .763 | -.028 | -.212 | .833 | -.067 | -.511 | .611 |
|  | Adulthood SES | -.067 | -.593 | .555 | -.040 | -.360 | .720 | -.046 | -.419 | .676 | -.067 | -.601 | .550 |
|  | Negative Life Events | .102 | .810 | .420 | -.039 | -.308 | .759 | .175 | 1.415 | .161 | .042 | .336 | .738 |

Bold values indicate significance at $p < 0.05$ ; findings did not survive FDR correction (0.05) for 7 tests. BNST, bed nucleus of the stria terminalis; PVN, paraventricular nucleus of the hypothalamus; sgACC, subgenual anterior cingulate cortex; SES, Socioeconomic Status; THQ, Trauma History Questionnaire.

### CVC Stressor-Evoked Activity and Affective Symptoms and Disorders

The amygdala showed a significant inverted U-shaped relationship with PTSS in the full model (β=-0.228, *P=*0.042, Figure 2A, Supplement, Table S6). The sgACC showed a linear relationship with PTSS at the threshold for significance in the full model (β=0.184, *P=*0.0498, Figure 2B, Supplement, Table S7).

**Figure 2.**
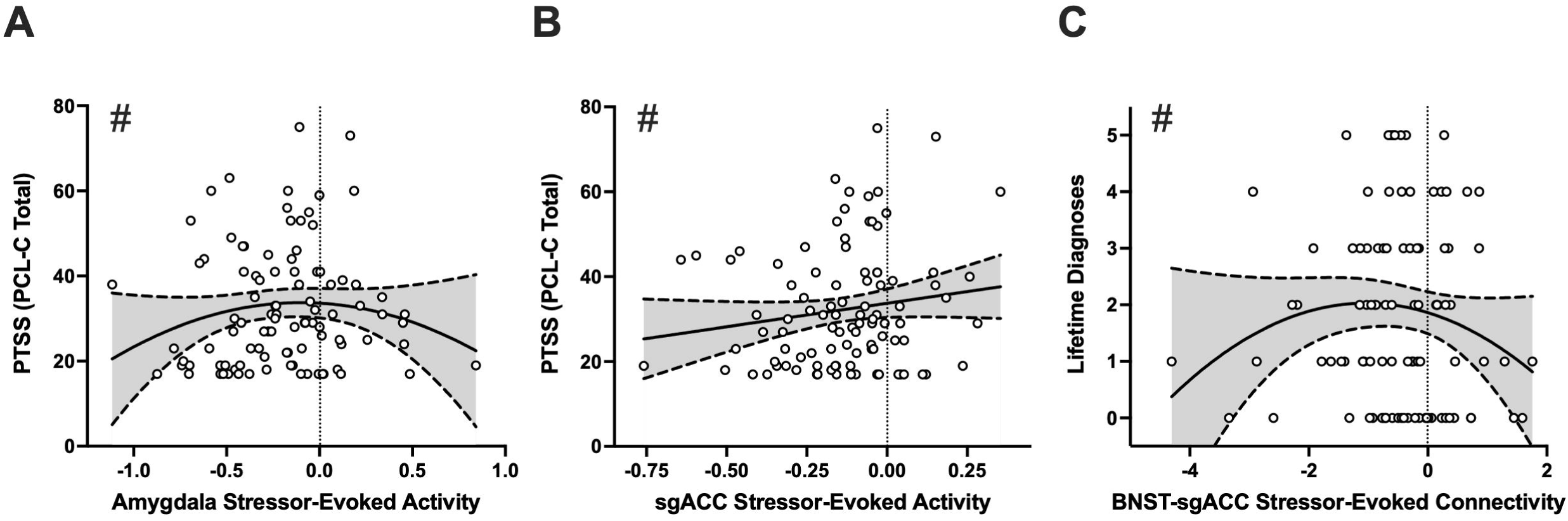
Central Visceral Circuit stressor-evoked activity and connectivity are associated with affective symptoms and diagnoses. **(A)** Amygdala activity showed a significant inverted U-shaped relationship with post-traumatic stress symptoms (PCL-C Total), with blunted reactivity (near-zero) associated with greater symptoms (β=-0.228, *P=*0.042). **(B)** sgACC activity showed a linear relationship with post-traumatic stress symptoms, such that lower reactivity (more relative activation) was associated with greater symptoms (β=0.184, *P=*0.0498). **(C)** BNST-sgACC connectivity showed a significant inverted U-shaped relationship with lifetime affective diagnoses, with diagnoses peaking at near-zero connectivity and fewer diagnoses toward both ends of the connectivity range (β=-0.285, *P=*0.035). #Indicates the association was significant in the full model with the adulthood covariates (traumatic events >18 years, education level/SES, past-year negative life events). (BNST, bed nucleus of the stria terminalis; sgACC, subgenual anterior cingulate cortex; PCL-C, PTSD Checklist–Civilian Version).

### CVC Stressor-Evoked Connectivity and Affective Symptoms and Disorders

BNST-sgACC connectivity showed a significant, inverted U-shaped relationship with the number of lifetime diagnoses (β=-0.285, *P=*0.035), remaining significant in the full model (β=-0.280, *P=*0.025, Figure 2C, Supplement, Table S8).

### CVC Stressor-Evoked Activity and Cardiovascular Outcomes

Stressor-evoked amygdala activity showed a significant relationship with DBP reactivity (β=0.272, *P=*0.011), remaining significant in the full model (β=0.286, *P=*0.009, Figure 3A). Stressor-evoked amygdala activity showed a significant relationship with DBP recovery (β=-0.277, *P=*0.006), remaining significant in the full model (β=-0.304, *P*=0.003, Figure 3B, Table S9).

**Figure 3.**
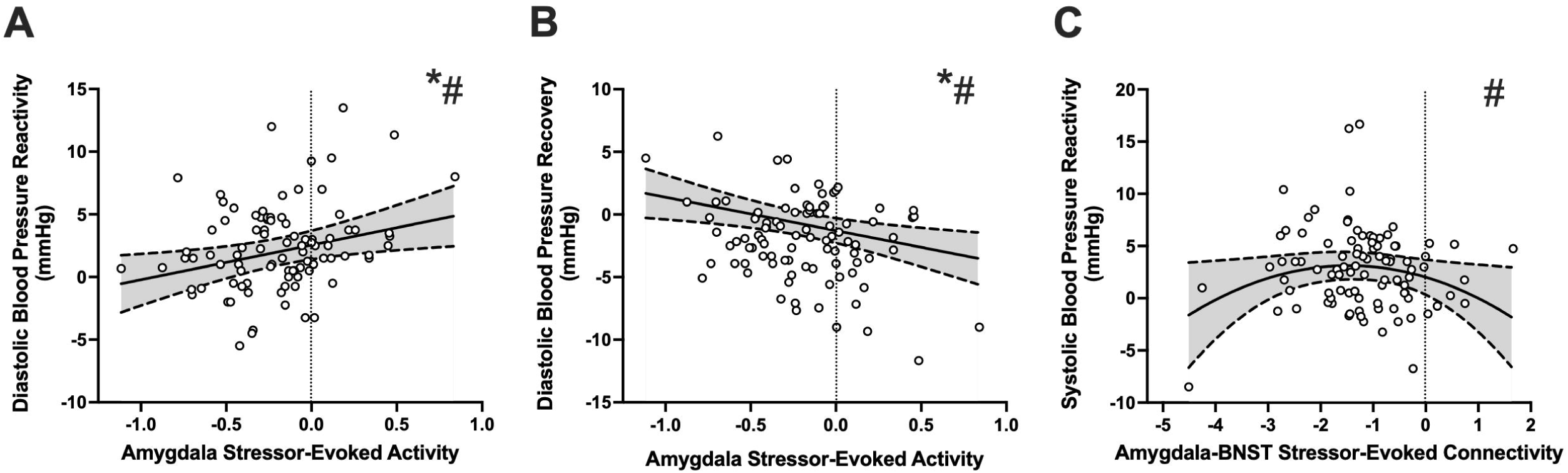
Amygdala stressor-evoked activity and connectivity are associated with cardiovascular stress reactivity. **(A)** Amygdala activity was linearly associated with diastolic blood pressure (DBP) reactivity, with greater deactivation (reactivity) associated with blunted DBP reactivity (β=0.272, *P=*0.011). **(B)** Amygdala activity was linearly associated with DBP recovery, with greater reactivity associated with less recovery (β=- 0.277, *P=*0.006). **(C)** Amygdala-BNST connectivity showed a significant inverted U-shaped relationship with systolic blood pressure (SBP) reactivity, with SBP reactivity greatest at moderate connectivity and lower toward both ends of the connectivity range (β=-0.458, *P=*0.028). *Indicates survival of FDR correction (0.05, for three tests, baseline, stress, and recovery for DBP and SBP separately). #Indicates the association was significant in the full model with the adulthood covariates (traumatic events >18 years, education level/SES, past-year negative life events). (BNST, bed nucleus of the stria terminalis; mmHg, millimeters of Mercury).

### CVC Stressor-Evoked Connectivity and Cardiovascular Outcomes

Stressor-evoked Amygdala-BNST connectivity showed a significant inverted U-shaped relationship with SBP reactivity (β=-0.458, *P=*0.028), remaining significant in the full model (β=-0.514, *P*=0.018, Figure 3C, Table S10). There was a linear trend between BNST-sgACC connectivity and baseline DBP in the full model (β=- 0.181, *P=*0.053, data not shown). (Potential outliers are reported in the Supplement, Table S11.)

## Discussion

Childhood adversity disrupts stress response systems, impacting mental health(114–116) and cardiovascular(117, 118) outcomes, though underlying neurobiological mechanisms remain unclear. We assessed a CVC as a candidate neural mechanism linking childhood adversity with affective symptoms/disorders and cardiovascular reactivity. The current study builds on evidence that CVC structure and function may be differentially impacted by threat and deprivation(88, 119), extending this work to stressor-evoked activity/connectivity. Our findings revealed two opposing curvilinear relationships: childhood threat with stressor-evoked amygdala activity (inverted U-shaped) and deprivation with sgACC activity (U-shaped), indicating potential threshold-mediated effects of adversity. These patterns partially supported our hypotheses; moderate threat was associated with blunted amygdala reactivity and moderate deprivation with heightened sgACC reactivity, with both relationships reversing with more severe adversity. Deprivation was also associated with stressor-evoked BNST connectivity, supporting a hub-like role for the BNST in integrating stress signals. CVC measures were, in turn, associated with affective and cardiovascular outcomes, suggesting that adversity-related CVC differences may contribute to stress-related mental and cardiovascular health trajectories.

Childhood threat (CTQ Threat or THQ 0-11) was associated with amygdala stressor-evoked activity, whereby low and high threat was associated with greater amygdala deactivation (i.e., *greater* stress reactivity; greater difference between stress and control conditions(86)) and moderate threat was associated with more blunted reactivity (i.e., less difference between task conditions) suggesting adaptive to maladaptive amygdala reactivity across a range of threat severity. This is consistent with studies linking maltreatment to heightened amygdala emotional reactivity(120–122) and stress reactivity, linearly, in a healthy sample using the same task(86). The curvilinear associations observed here may reflect the broader threat severity distribution in our transdiagnostic sample, indicating thresholds at which adaptive and maladaptive stress responses may occur.

A U-shaped relationship emerged between childhood deprivation (SED) and stressor-evoked sgACC activity, where low and high deprivation were associated with blunted sgACC stress reactivity (i.e., near zero, little difference between stress and control conditions) again suggesting adaptive to maladaptive reactivity across a range of severity. The sgACC is central in emotion regulation and negative affect, with neural alterations in mood disorders(62, 123). Further, the sgACC is itself visceromotor and facilitates regulatory control over CVC regions(124–126), namely amygdala and BNST. In individuals with a history of deprivation the sgACC may not mount an appropriate stress response, contributing to CVC dysregulation. Indeed, greater deprivation was also associated with weaker BNST-sgACC connectivity (i.e., near-zero, little difference in task-related coupling), which may impair sgACC ability to effectively modulate BNST stress responses and further downstream physiological effects.

All deprivation-related connectivity findings involved BNST, consistent with its hub-like role integrating autonomic and viscerosensory stress signals across CVCs(127–129), with greater deprivation linked to weaker BNST-related connectivity overall. Conversely, greater trauma was associated with greater PVN-BNST connectivity (i.e., more negative values indicate greater coupling during incongruent compared to congruent conditions). Given the inhibitory relationship between BNST and PVN and subsequent inhibitory influences on HPA responses(130–132), trauma may strengthen this connection, yielding less physiological stress reactivity. Our previous resting-state findings demonstrated that greater trauma was associated with lower BNST-PVN connectivity(87); current findings are consistent with our prior hypothesis that childhood threat may prime resting-state CVC connectivity to more actively engage direct connections (between BNST and PVN) during stress. These findings support a growing body of literature indicating that threat and deprivation distinctly influence neural structure and function to shape downstream mental and physical health outcomes(119, 133–135).

Analyses revealed stressor-evoked CVC associations with affective outcomes; amygdala and sgACC stressor-evoked activity were linked to PTSS (curvilinearly and linearly, respectively). Individuals with greater amygdala reactivity (deactivation *or* activation) had fewer PTSS, with blunted reactivity associated with higher symptom severity; this suggests that the amygdala’s capacity to mount an appropriate stress response is adaptive for clinical outcomes. These findings extend prior reports of amygdala hyperresponsivity to trauma-related stimuli in PTSD(136) to responses under mild cognitive stress in a continuous adversity-exposed sample. Greater sgACC reactivity was associated with fewer PTSS, also suggesting that adequate sgACC reactivity is adaptive. This is congruent with structural findings linking less sgACC volume with greater PTSS(137, 138), potentially reflecting decreased capacity for stress and emotion regulation.

An inverted U-shaped relationship also emerged between BNST-sgACC stressor-evoked connectivity and lifetime affective diagnoses, with diagnoses peaking at near-zero (weak) connectivity. This is consistent with prior studies demonstrating extensive sgACC-BNST connectivity(51, 53, 139), and linking lower sgACC- limbic connectivity to heightened depressive and PTSD symptoms(140, 141). Given sgACC’s role in emotion regulation(62) and large-scale network integration(142), disrupted BNST-sgACC connectivity may compromise regulation of stress-related signaling and coordination across CVCs, increasing vulnerability to psychopathology, whereas stronger connectivity may be protective.

Our findings linked stressor-evoked CVC activity/connectivity with cardiovascular outcomes; greater amygdala reactivity was associated with blunted DBP reactivity, consistent with previous work in a smaller healthy young-adult sample showing that greater amygdala deactivation in response to a similar stress task was associated with blunted mean arterial pressure reactivity(143). Greater amygdala reactivity was also associated with less DBP recovery, together suggesting that individuals with greater amygdala reactivity may display less dynamic/flattened autonomic responsiveness.

Additionally, we observed an inverted U-shaped relationship between stressor-evoked amygdala-BNST connectivity and SBP reactivity, greatest at moderate connectivity, and lower with greater connectivity at both connectivity extremes (more incongruent-dominant or more congruent-dominant coupling). The amygdala and BNST are reciprocally connected nodes(49, 144) that send convergent, predominantly GABAergic projections to hypothalamic and brainstem circuits regulating autonomic outflow(50, 145–148). Both modulate brainstem baroreflex circuitry(73, 107, 149), biasing autonomic control of the heart and vasculature. Given this predominantly inhibitory output, depending on local targets, more coordinated amygdala–BNST coupling during stress may yield sympathoinhibition and an attenuated pressor response.

Prior work has documented links between childhood adversity and dysregulated stress reactivity, with blunted stress-related cardiovascular psychophysiology reported in healthy samples(29, 150, 151). In our abuse-enriched, transdiagnostic sample, *greater* amygdala reactivity was associated with blunted cardiovascular (i.e., DBP) stress reactivity and, consistent with the curvilinear relationship described above, with more severe abuse. Our findings may support a model in which childhood threat-related alterations in amygdala reactivity contribute to blunted cardiovascular reactivity. Given that blunted cardiovascular reactivity is associated with adverse health outcomes(152–154), our findings may have clinical implications for cardiovascular health. A large, multivariate study demonstrated that lower levels of amygdala emotional reactivity, as part of a distributed corticolimbic system, were associated with greater carotid artery intima-media thickness, a marker of preclinical atherosclerosis(155). Although the paradigms differ, these findings situate amygdala function within a broader system that may contribute risk to cardiovascular disease in the context of adversity.

Limitations of this work, namely retrospective reporting and cross-sectional design, have been discussed previously, and are mitigated by our sample’s relatively uniform distribution across childhood physical abuse severity. While potential CVC links were identified between childhood adversity and affective and cardiovascular outcomes, our study is underpowered to test mediation directly. Further, with the absence of neuroendocrine measures, we are unable to address links between adversity, CVCs and neuroendocrine reactivity. Additionally, current resolution at 3 Tesla may limit detection of existing relationships between childhood adversity and BNST and PVN stressor-evoked activity; larger studies using ultra-high field approaches may be necessary to examine these relationships.

This study revealed significant relationships between childhood adversity and stressor-evoked activity within the amygdala and sgACC in relation to threat and deprivation, respectively, as well as a linear relationship between deprivation and BNST-sgACC connectivity. The curvilinear nature of the amygdala and sgACC findings suggests thresholds at which vulnerabilities to psychopathology and dysregulated stress reactivity may emerge or be exacerbated. These adversity-related continua of reactivity were, in turn, associated with outcomes indicating differential risk: blunted amygdala and lower sgACC reactivity, along with weaker BNST-sgACC regulatory signaling, may confer risk for psychopathology, whereas heightened amygdala reactivity may confer risk for dysregulated cardiovascular stress responses. Future research examining dimensions of adversity along with CVC subnuclei-specific relationships using higher resolution imaging may inform targeted interventions designed to mitigate the long-term negative health consequences of childhood adversity.

## Data Availability

All data produced in the present study are available upon reasonable request to the corresponding author.

## Acknowledgements

We thank Hussain M. Alkhars for analyses of subjective task ratings, Shannon Gallagher for preliminary work related to calculating cardiovascular reactivity, Anne Germain for providing infrastructure and facilitating participant recruitment, Mark Jones for clinical supervision, Noelle Rode for database construction and Peter J. Gianaros for providing comments on an earlier draft of this manuscript.

## AI Disclosure Statement

AI assisted in copy editing human-written language to find typographical or grammatical errors and provide suggestions for condensing language for length. No generative AI was used to generate scientific content, conduct analyses, create or alter figures, or draw scientific conclusions.

## References

1. Sheridan MA, McLaughlin KA (2014): Dimensions of early experience and neural development: deprivation and threat. Trends in Cognitive Sciences. 18:580–585.

2. McLaughlin KA, Sheridan MA, Lambert HK (2014): Childhood adversity and neural development: Deprivation and threat as distinct dimensions of early experience. Neuroscience & Biobehavioral Reviews. 47:578–591.

3. Fani N, Bradley-Davino B, Ressler KJ, McClure-Tone EB (2010): Attention Bias in Adult Survivors of Childhood Maltreatment with and without Posttraumatic Stress Disorder. Cogn Ther Res. 35:57–67.

4. McCrory EJ, De Brito SA, Kelly PA, Bird G, Sebastian CL, Mechelli A, et al. (2013): Amygdala activation in maltreated children during pre-attentive emotional processing. The British Journal of Psychiatry. 202:269–276.

5. van Harmelen AL, van Tol MJ, Demenescu LR, van der Wee NJ, Veltman DJ, Aleman A, et al. (2013): Enhanced amygdala reactivity to emotional faces in adults reporting childhood emotional maltreatment. Soc Cogn Affect Neurosci. 8:362–369.

6. Teicher MH, Samson JA (2016): Annual Research Review: Enduring neurobiological effects of childhood abuse and neglect. Journal of Child Psychology and Psychiatry. 57:241–266.

7. Teicher MH, Samson JA, Anderson CM, Ohashi K (2016): The effects of childhood maltreatment on brain structure, function and connectivity. Nature Reviews Neuroscience. 17:652–666.

8. Hillis S, Mercy J, Amobi A, Kress H (2016): Global Prevalence of Past-year Violence Against Children: A Systematic Review and Minimum Estimates. Pediatrics. 137:e20154079.

9. Cuartas J, McCoy DC, Rey-Guerra C, Britto PR, Beatriz E, Salhi C (2019): Early childhood exposure to non-violent discipline and physical and psychological aggression in low- and middle-income countries: National, regional, and global prevalence estimates. Child Abuse & Neglect. 92:93–105.

10. Hoppen TH, Chalder T (2018): Childhood adversity as a transdiagnostic risk factor for affective disorders in adulthood: A systematic review focusing on biopsychosocial moderating and mediating variables. Clinical Psychology Review. 65:81–151.

11. Juwariah T, Suhariadi F, Soedirham O, Priyanto A, Setiyorini E, Siskaningrum A, et al. (2022): Childhood adversities and mental health problems: A systematic review. Journal of public health research. 11:22799036221106613.

12. Rozanski S, Schmidt A, John A, Gaysina D (2021): Childhood neglect and trajectories of affective symptoms throughout adulthood: A British birth cohort study. Journal of Affective Disorders. 295:416–421.

13. Teicher MH, Samson JA (2013): Childhood maltreatment and psychopathology: a case for ecophenotypic variants as clinically and neurobiologically distinct subtypes. American journal of psychiatry. 170:1114–1133.

14. Teicher MH, Ohashi K, Khan A (2020): Additional Insights into the Relationship Between Brain Network Architecture and Susceptibility and Resilience to the Psychiatric Sequelae of Childhood Maltreatment. Adversity Resil Sci. 1:49–64.

15. Barr DA (2020): Stepping Stones From Childhood Adversity to Cardiovascular Disease and Premature Mortality. Journal of the American Heart Association. 9:e016162.

16. Bengtsson J, Elsenburg LK, Andersen GS, Larsen ML, Rieckmann A, Rod NH (2023): Childhood adversity and cardiovascular disease in early adulthood: a Danish cohort study. European Heart Journal. 44:586–593.

17. Suglia SF, Koenen KC, Boynton-Jarrett R, Chan PS, Clark CJ, Danese A, et al. (2018): Childhood and Adolescent Adversity and Cardiometabolic Outcomes: A Scientific Statement From the American Heart Association. Circulation. 137:e15–e28.

18. Danese A, van Harmelen A-L (2017): The hidden wounds of childhood trauma. European Journal of Psychotraumatology. 8:1375840.

19. Nelson CA, Bhutta ZA, Burke Harris N, Danese A, Samara M (2020): Adversity in childhood is linked to mental and physical health throughout life. BMJ. 371:m3048.

20. Heim C, Newport DJ, Bonsall R, Miller AH, Nemeroff CB (2001): Altered pituitary-adrenal axis responses to provocative challenge tests in adult survivors of childhood abuse. American Journal of Psychiatry. 158:575–581.

21. Heim C, Newport DJ, Heit S, Graham YP, Wilcox M, Bonsall R, et al. (2000): Pituitary-adrenal and autonomic responses to stress in woman after sexual and physical abuse in childhood. JAMA. 284:592–597.

22. Heim C, Newport DJ, Mletzko T, Miller AH, Nemeroff CB (2008): The link between childhood trauma and depression: insights from HPA axis studies in humans. Psychoneuroendocrinology. 33:693–710.

23. Peckins MK, Roberts AG, Hein TC, Hyde LW, Mitchell C, Brooks-Gunn J, et al. (2020): Violence exposure and social deprivation is associated with cortisol reactivity in urban adolescents. Psychoneuroendocrinology. 111:104426.

24. Bourassa KJ, Moffitt TE, Harrington H, Houts R, Poulton R, Ramrakha S, et al. (2020): Lower Cardiovascular Reactivity Is Associated With More Childhood Adversity and Poorer Midlife Health: Replicated Findings From the Dunedin and MIDUS Cohorts. Clin Psychol Sci. 9:961–978.

25. Brindle RC, Pearson A, Ginty AT (2022): Adverse childhood experiences (ACEs) relate to blunted cardiovascular and cortisol reactivity to acute laboratory stress: A systematic review and meta-analysis. Neuroscience & Biobehavioral Reviews. 134:104530.

26. Dempster KS, O’Leary DD, MacNeil AJ, Hodges GJ, Wade TJ (2021): Linking the hemodynamic consequences of adverse childhood experiences to an altered HPA axis and acute stress response. *Brain*, Behavior, and Immunity. 93:254–263.

27. Keogh TM, Howard S, Gallagher S (2022): Early life adversity and blunted cardiovascular reactivity to acute psychological stress: The role of current depressive symptoms. Biopsychosocial Science and Medicine. 84:170–178.

28. Keogh TM, Howard S, Gallagher S, Ginty AT (2023): Cluster analysis reveals distinct patterns of childhood adversity, behavioral disengagement, and depression that predict blunted heart rate reactivity to acute psychological stress. Annals of Behavioral Medicine. 57:61–73.

29. Lovallo WR, Farag NH, Sorocco KH, Cohoon AJ, Vincent AS (2012): Lifetime Adversity Leads to Blunted Stress Axis Reactivity: Studies from the Oklahoma Family Health Patterns Project. Biological Psychiatry. 71:344–349.

30. Carpenter L, Shattuck T, Tyrka A, Geracioti T, Price L (2011): Effect of childhood physical abuse on cortisol stress response. Psychopharmacology. 214:367–375.

31. Carpenter LL, Carvalho JP, Tyrka AR, Wier LM, Mello AF, Mello MF, et al. (2007): Decreased adrenocorticotropic hormone and cortisol responses to stress in healthy adults reporting significant childhood maltreatment. Biological Psychiatry. 62:1080–1087.

32. Peckins MK, Susman EJ, Negriff S, Noll J, Trickett PK (2015): Cortisol profiles: A test for adaptive calibration of the stress response system in maltreated and nonmaltreated youth. Development and Psychopathology. 27:1461–1470.

33. Doom JR, Cicchetti D, Rogosch FA (2014): Longitudinal Patterns of Cortisol Regulation Differ in Maltreated and Nonmaltreated Children. Journal of the American Academy of Child & Adolescent Psychiatry. 53:1206–1215.

34. Bernard K, Frost A, Bennett CB, Lindhiem O (2017): Maltreatment and diurnal cortisol regulation: A meta-analysis. Psychoneuroendocrinology. 78:57–67.

35. Chen E, Cohen S, Miller GE (2009): How Low Socioeconomic Status Affects 2-Year Hormonal Trajectories in Children. Psychological Science. 21:31–37.

36. Lupien SJ, King S, Meaney MJ, McEwen BS (2001): Can poverty get under your skin? Basal cortisol levels and cognitive function in children from low and high socioeconomic status. Development and Psychopathology. 13:653–676.

37. Cohen S, Doyle W, Baum A (2006): Socioeconomic Status Is Associated With Stress Hormones. Psychosomatic Medicine. 68:414–420.

38. Le-Scherban F, Brenner AB, Hicken MT, Needham BL, Seeman T, Sloan RP, et al. (2018): Child and Adult Socioeconomic Status and the Cortisol Response to Acute Stress: Evidence From the Multi-Ethnic Study of Atherosclerosis. Psychosom Med. 80:184–192.

39. Del Giudice M, Ellis BJ, Shirtcliff EA (2011): The Adaptive Calibration Model of stress responsivity. Neuroscience and biobehavioral reviews. 35:1562–1592.

40. Rinaman L, Banihashemi L, Koehnle TJ (2011): Early life experience shapes the functional organization of stress-responsive visceral circuits. Physiol Behav. 104:632–640.

41. Luiten PGM, Ter Horst GJ, Karst H, Steffens AB (1985): The course of paraventricular hypothalamic efferents to autonomic structures in medulla and spinal cord. Brain Research. 329:374–378.

42. Herman JP, Cullinan WE, Ziegler DR, Tasker JG (2002): Role of the paraventricular nucleus microenvironment in stress integration. European Journal of Neuroscience. 16:381–385.

43. Lozić M, Šarenac O, Murphy D, Japundžić-Žigon N (2018): Vasopressin, Central Autonomic Control and Blood Pressure Regulation. Current Hypertension Reports. 20:11.

44. Feetham CH, O’Brien F, Barrett-Jolley R (2018): Ion Channels in the Paraventricular Hypothalamic Nucleus (PVN); Emerging Diversity and Functional Roles. Front Physiol. Volume 9.

45. Povysheva N, Zheng H, Rinaman L (2021): Glucagon-like peptide 1 receptor-mediated stimulation of a GABAergic projection from the bed nucleus of the stria terminalis to the hypothalamic paraventricular nucleus. Neurobiology Stress. 15:100363.

46. Prewitt CM, Herman JP (1994): Lesion of the Central Nucleus of the Amygdala Decreases Basal CRH mRNA Expression and Stress-Induced ACTH Release. Annals of the New York Academy of Sciences. 746:438–440.

47. Chiou R-J, Kuo C-C, Yen C-T (2014): Comparisons of terminal densities of cardiovascular function-related projections from the amygdala subnuclei. Autonomic Neuroscience. 181:21–30.

48. Bienkowski MS, Wendel ES, Rinaman L (2013): Organization of multisynaptic circuits within and between the medial and the central extended amygdala. J Comp Neurol. 521:3406–3431.

49. Dong H-W, Petrovich GD, Swanson LW (2001): Topography of projections from amygdala to bed nuclei of the stria terminalis. Brain Research Reviews. 38:192–246.

50. Dong H-W, Petrovich GD, Watts AG, Swanson LW (2001): Basic organization of projections from the oval and fusiform nuclei of the bed nuclei of the stria terminalis in adult rat brain. J Comp Neurol. 436:430–455.

51. Freedman LJ, Insel TR, Smith Y (2000): Subcortical projections of area 25 (subgenual cortex) of the macaque monkey. J Comp Neurol. 421:172–188.

52. Vertes RP (2004): Differential projections of the infralimbic and prelimbic cortex in the rat. Synapse. 51:32–58.

53. Ongur D, Price JL (2000): The organization of networks within the orbital and medial prefrontal cortex of rats, monkeys and humans. Cerebral Cortex. 10:206.

54. Sharma KK, Kelly EA, Pfeifer CW, Fudge JL (2019): Translating Fear Circuitry: Amygdala Projections to Subgenual and Perigenual Anterior Cingulate in the Macaque. Cerebral Cortex. 30:550–562.

55. Alexander L, Clarke HF, Roberts AC (2019): A Focus on the Functions of Area 25. Brain Sci. 9:129.

56. Avery SN, Clauss JA, Blackford JU (2016): The Human BNST: Functional Role in Anxiety and Addiction. Neuropsychopharmacology. 41:126–141.

57. Avery SN, Clauss JA, Winder DG, Woodward N, Heckers S, Blackford JU (2014): BNST neurocircuitry in humans. NeuroImage. 91:311–323.

58. Somerville LH, Whalen PJ, Kelley WM (2010): Human Bed Nucleus of the Stria Terminalis Indexes Hypervigilant Threat Monitoring. BPS. 68:416–424.

59. Herrmann MJ, Boehme S, Becker MPI, Tupak SV, Guhn A, Schmidt B, et al. (2016): Phasic and sustained brain responses in the amygdala and the bed nucleus of the stria terminalis during threat anticipation. Human Brain Mapping. 37:1091–1102.

60. Fendt M, Siegl S, Steiniger-Brach B (2005): Noradrenaline transmission within the ventral bed nucleus of the stria terminalis is critical for fear behavior induced by trimethylthiazoline, a component of fox odor. The Journal of Neuroscience. 25:5998–6004.

61. Schweimer J, Fendt M, Schnitzler H-U (2005): Effects of clonidine injections into the bed nucleus of the stria terminalis on fear and anxiety behavior in rats. European Journal of Pharmacology. 507:117–124.

62. Drevets WC, Savitz J, Trimble M (2008): The subgenual anterior cingulate cortex in mood disorders. CNS spectrums. 13:663.

63. Gotlib IH, Sivers H, Gabrieli JDE, Whitfield-Gabrieli S, Goldin P, Minor KL, et al. (2005): Subgenual anterior cingulate activation to valenced emotional stimuli in major depression. Neuroreport. 16:1731.

64. Lebow MA, Chen A (2016): Overshadowed by the amygdala: the bed nucleus of the stria terminalis emerges as key to psychiatric disorders. Nature Publishing Group. 21:450–463.

65. Thayer JF, Lane RD (2000): A model of neurovisceral integration in emotion regulation and dysregulation. Journal of Affective Disorders. 61:201–216.

66. Clauss J (2019): Extending the neurocircuitry of behavioural inhibition: a role for the bed nucleus of the stria terminalis in risk for anxiety disorders. Gen Psychiatry. 32:e100137.

67. Clauss JA, Avery SN, Benningfield MM, Blackford JU (2019): Social anxiety is associated with BNST response to unpredictability. Depression and Anxiety. 36:666–675.

68. Jin S, Liu W, Hu Y, Liu Z, Xia Y, Zhang X, et al. (2023): Aberrant functional connectivity of the bed nucleus of the stria terminalis and its age dependence in children and adolescents with social anxiety disorder. Asian Journal of Psychiatry. 82:103498.

69. Feola B, Flook EA, Gardner H, Phan KL, Gwirtsman H, Olatunji B, et al. (2023): Altered bed nucleus of the stria terminalis and amygdala responses to threat in combat veterans with posttraumatic stress disorder. Journal of Traumatic Stress. 36:359–372.

70. Peng Y, Knotts JD, Young KS, Bookheimer SY, Nusslock R, Zinbarg RE, et al. (2023): Threat Neurocircuitry Predicts the Development of Anxiety and Depression Symptoms in a Longitudinal Study. Biological Psychiatry: Cognitive Neuroscience and Neuroimaging. 8:102–110.

71. Slabe Z, Balesar RA, Verwer RWH, Van Heerikhuize JJ, Pechler GA, Zorović M, et al. (2023): Alterations in pituitary adenylate cyclase-activating polypeptide in major depressive disorder, bipolar disorder, and comorbid depression in Alzheimer’s disease in the human hypothalamus and prefrontal cortex. Psychological Medicine. 53:7537–7549.

72. Alexander L, Wood CM, Gaskin PLR, Sawiak SJ, Fryer TD, Hong YT, et al. (2020): Over-activation of primate subgenual cingulate cortex enhances the cardiovascular, behavioral and neural responses to threat. Nat Commun. 11:5386.

73. Crestani CC, Alves FHF, Gomes FV, Resstel LBM, Correa FMA, Herman JP (2013): Mechanisms in the Bed Nucleus of the Stria Terminalis Involved in Control of Autonomic and Neuroendocrine Functions: A Review. Curr Neuropharmacol. 11:141–159.

74. Savić B, Murphy D, Japundžić-Žigon N (2022): The Paraventricular Nucleus of the Hypothalamus in Control of Blood Pressure and Blood Pressure Variability. Front Physiol. Volume 13–2022.

75. Coote JH (2004): A role for the paraventricular nucleus of the hypothalamus in the autonomic control of heart and kidney. Experimental Physiology. 90:169–173.

76. Dampney RA, Michelini LC, Li D-P, Pan H-L (2018): Regulation of sympathetic vasomotor activity by the hypothalamic paraventricular nucleus in normotensive and hypertensive states. Am J Physiol-heart C. 315:H1200–H1214.

77. Schwaber JS, Kapp BS, Higgins GA, Rapp PR (1982): Amygdaloid and basal forebrain direct conections with the nucleus of the solitary tract and the dorsal motor nucleus. The Journal of Neuroscience. 2:1424–1438.

78. Card JP, Levitt P, Gluhovsky M, Rinaman L (2005): Early experience modifies the postnatal assembly of autonomic emotional motor circuits in rats. The Journal of Neuroscience. 25:9102–9111.

79. Banihashemi L, Rinaman L (2010): Repeated brief postnatal maternal separation enhances hypothalamic gastric autonomic circuits in juvenile rats. Neuroscience. 165:265–277.

80. Banihashemi L, O’Neill EJ, Rinaman L (2011): Central neural responses to restraint stress are altered in rats with an early life history of repeated brief maternal separation. Neuroscience. 192:413–428.

81. Klumpers F, Kroes MCW, Baas JMP, Fernández G (2017): How Human Amygdala and Bed Nucleus of the Stria Terminalis May Drive Distinct Defensive Responses. The Journal of Neuroscience. 37:9645.

82. Zhong X, Ming Q, Dong D, Sun X, Cheng C, Xiong G, et al. (2019): Childhood Maltreatment Experience Influences Neural Response to Psychosocial Stress in Adults: An fMRI Study. Front Psychol. 10.

83. Banihashemi L, Wallace ML, Peng CW, Stinley MM, Germain A, Herringa RJ (2020): Interactions between childhood maltreatment and combat exposure trauma on stress-related activity within the cingulate cortex: a pilot study. Mil Psychol. 32:176–185.

84. Dedovic K, D’Aguiar C, Pruessner JC (2009): What Stress Does to Your Brain: A Review of Neuroimaging Studies. The Canadian Journal of Psychiatry. 54:6–15.

85. Steptoe A, Hamer M, Lin J, Blackburn EH, Erusalimsky JD (2017): The longitudinal relationship between cortisol responses to mental stress and leukocyte telomere attrition. The Journal of Clinical Endocrinology & Metabolism. 102:962–969.

86. Banihashemi L, Sheu LK, Midei AJ, Gianaros PJ (2015): Childhood physical abuse predicts stressor-evoked activity within central visceral control regions. Soc Cogn Affect Neurosci. 10:474–485.

87. Banihashemi L, Peng CW, Rangarajan A, Karim HT, Wallace ML, Sibbach BM, et al. (2022): Childhood Threat Is Associated With Lower Resting-State Connectivity Within a Central Visceral Network. Front Psychol. 13:805049.

88. Banihashemi L, Peng CW, Verstynen T, Wallace ML, Lamont DN, Alkhars HM, et al. (2021): Opposing relationships of childhood threat and deprivation with stria terminalis white matter. Human Brain Mapping. 42:2445–2460.

89. Rudolph KD, Davis MM, Skymba HV, Modi HH, Telzer EH (2021): Social experience calibrates neural sensitivity to social feedback during adolescence: A functional connectivity approach. Developmental Cognitive Neuroscience. 47:100903.

90. Oshri A, Howard CJ, Zhang L, Reck A, Cui Z, Liu S, et al. (2024): Strengthening through adversity: The hormesis model in developmental psychopathology. Development and psychopathology. 36:2390–2406.

91. Harnett NG, Wheelock MD, Wood KH, Ladnier JC, Mrug S, Knight DC (2015): Affective state and locus of control modulate the neural response to threat. NeuroImage. 121:217–226.

92. Wood KH, Ver Hoef LW, Knight DC (2014): The amygdala mediates the emotional modulation of threat-elicited skin conductance response. Emotion. 14:693–700.

93. Bernstein DP, Fink L, Handelsman L, Foote J, Lovejoy M, Wenzel K, et al. (1994): Initial reliability and validity of a new retrospective measure of child abuse and neglect. The American journal of psychiatry. 151:1132.

94. Green BL (1996): Trauma History Questionnaire. In: Stamm BH, editor. Measurement of stress, trauma, and adaptation. Lutherville, MD: The Sidran Press, pp 366–369.

95. Insana SP, Kolko DJ, Germain A (2012): Early-life trauma is associated with rapid eye movement sleep fragmentation among military veterans. Biol Psychol. 89:570–579.

96. Morris G, Berk M, Maes M, Carvalho AF, Puri BK (2019): Socioeconomic Deprivation, Adverse Childhood Experiences and Medical Disorders in Adulthood: Mechanisms and Associations. Molecular Neurobiology. 56:5866–5890.

97. Webb S, Janus M, Duku E, Raos R, Brownell M, Forer B, et al. (2017): Neighbourhood socioeconomic status indices and early childhood development. SSM - Population Health. 3:48–56.

98. Berti C, Pivetti M (2019): Childhood economic disadvantage and antisocial behavior: Intervening factors and pathways. Children and Youth Services Review. 97:120–126.

99. Reiss F (2013): Socioeconomic inequalities and mental health problems in children and adolescents: A systematic review. Social Science & Medicine. 90:24–31.

100. Ursache A, Merz EC, Melvin S, Meyer J, Noble KG (2017): Socioeconomic status, hair cortisol and internalizing symptoms in parents and children. Psychoneuroendocrinology. 78:142–150.

101. Winkleby MA, Jatulis DE, Frank E, Fortmann SP (1992): Socioeconomic status and health: how education, income, and occupation contribute to risk factors for cardiovascular disease. American Journal of Public Health. 82:816–820.

102. Cohen S, Tyrrell DAJ, Smith AP (1991): Psychological Stress and Susceptibility to the Common Cold. New England Journal of Medicine. 325:606–612.

103. Cohen S, Kamarck T, Mermelstein R (1983): A global measure of perceived stress. J Health Soc Behav. 24:385–396.

104. Spielberger CD, Gorsuch RL, Lushene R, Vagg PR, Jacobs GA (1983): Manual for the state-trait anxiety inventory (Palo Alto, CA, Consulting Psychologists Press). Inc.

105. McCrae RR, Costa Jr PT (2007): Brief versions of the NEO-PI-3. Journal of individual differences. 28:116.

106. Gianaros PJ, Sheu LK, Remo AM, Christie IC, Critchley HD, Wang J (2009): Heightened resting neural activity predicts exaggerated stressor-evoked blood pressure reactivity. Hypertension. 53:819–825.

107. Gianaros PJ, Onyewuenyi IC, Sheu LK, Christie IC, Critchley HD (2011): Brain systems for baroreflex suppression during stress in humans. Human Brain Mapping. 33:1700–1716.

108. Sheu LK, Jennings JR, Gianaros PJ (2012): Test-retest reliability of an fMRI paradigm for studies of cardiovascular reactivity. Psychophysiology. 49:873–884.

109. McLaren DG, Ries ML, Xu G, Johnson SC (2012): A generalized form of context-dependent psychophysiological interactions (gPPI): A comparison to standard approaches. NeuroImage. 61:1277–1286.

110. Beck AT, Steer RA, Ball R, Ranieri WF (1996): Comparison of Beck Depression Inventories-IA and-II in psychiatric outpatients. Journal of personality assessment. 67:588–597.

111. Wilkins KC, Lang AJ, Norman SB (2011): Synthesis of the psychometric properties of the PTSD checklist (PCL) military, civilian, and specific versions. Depression and anxiety. 28:596–606.

112. Lawson GM, Camins JS, Wisse L, Wu J, Duda JT, Cook PA, et al. (2017): Childhood socioeconomic status and childhood maltreatment: Distinct associations with brain structure. PLoS ONE. 12:e0175690–0175616.

113. Benjamini Y, Hochberg Y (1995): Controlling the False Discovery Rate: A Practical and Powerful Approach to Multiple Testing. Journal of the Royal Statistical Society Series B (Methodological). 57:289–300.

114. Hoppen TH, Chalder T (2018): Childhood adversity as a transdiagnostic risk factor for affective disorders in adulthood: A systematic review focusing on biopsychosocial moderating and mediating variables. Clin Psychol Rev. 65:81–151.

115. Juwariah T, Suhariadi F, Soedirham O, Priyanto A, Setiyorini E, Siskaningrum A, et al. (2022): Childhood adversities and mental health problems: A systematic review. J Public Health Res. 11:22799036221106613.

116. Rozanski S, Schmidt A, John A, Gaysina D (2021): Childhood neglect and trajectories of affective symptoms throughout adulthood: A British birth cohort study. J Affect Disord. 295:416–421.

117. Danese A, van Harmelen AL (2017): The hidden wounds of childhood trauma. Eur J Psychotraumatol. 8:137584.

118. Nelson CA, Scott RD, Bhutta ZA, Harris NB, Danese A, Samara M (2020): Adversity in childhood is linked to mental and physical health throughout life. Bmj. 371:m3048.

119. Banihashemi L, Peng CW, Verstynen T, Wallace ML, Lamont DN, Alkhars HM, et al. (2021): Opposing relationships of childhood threat and deprivation with stria terminalis white matter. Human Brain Mapping. n/a.

120. Hein TC, Monk CS (2017): Research Review: Neural response to threat in children, adolescents, and adults after child maltreatment – a quantitative meta-analysis. Journal of Child Psychology and Psychiatry. 58:222–230.

121. Kuo P-C, Yao Z-F (2025): Amygdala hyperactivation in childhood maltreatment: An ALE-based meta-analysis on emotion-related processing. Neuroscience & Biobehavioral Reviews. 174:106180.

122. Puetz VB, Viding E, Gerin MI, Pingault JB, Sethi A, Knodt AR, et al. (2020): Investigating patterns of neural response associated with childhood abuse v. childhood neglect. Psychol Med. 50:1398–1407.

123. Phillips ML, Ladouceur CD, Drevets WC (2008): A neural model of voluntary and automatic emotion regulation: implications for understanding the pathophysiology and neurodevelopment of bipolar disorder. Molecular Psychiatry. 13:833–857.

124. Alexander L, Wood CM, Gaskin PLR, Sawiak SJ, Fryer TD, Hong YT, et al. (2020): Over-activation of primate subgenual cingulate cortex enhances the cardiovascular, behavioral and neural responses to threat. Nat Commun. 11:5386.

125. Sharma KK, Kelly EA, Pfeifer CW, Fudge JL (2020): Translating Fear Circuitry: Amygdala Projections to Subgenual and Perigenual Anterior Cingulate in the Macaque. Cereb Cortex. 30:550–562.

126. Tillman RM, Stockbridge MD, Nacewicz BM, Torrisi S, Fox AS, Smith JF, et al. (2018): Intrinsic functional connectivity of the central extended amygdala. Hum Brain Mapp. 39:1291–1312.

127. Dong HW, Petrovich GD, Watts AG, Swanson LW (2001): Basic organization of projections from the oval and fusiform nuclei of the bed nuclei of the stria terminalis in adult rat brain. J Comp Neurol. 436:430–455.

128. Kim SY, Adhikari A, Lee SY, Marshel JH, Kim CK, Mallory CS, et al. (2013): Diverging neural pathways assemble a behavioural state from separable features in anxiety. Nature. 496:219–223.

129. Ch’ng S, Fu J, Brown RM, McDougall SJ, Lawrence AJ (2018): The intersection of stress and reward: BNST modulation of aversive and appetitive states. Prog Neuropsychopharmacol Biol Psychiatry. 87:108–125.

130. Radley JJ, Arias CM, Sawchenko PE (2006): Regional Differentiation of the Medial Prefrontal Cortex in Regulating Adaptive Responses to Acute Emotional Stress. J Neurosci. 26:12967–12976.

131. Radley JJ, Gosselink KL, Sawchenko PE (2009): A Discrete GABAergic Relay Mediates Medial Prefrontal Cortical Inhibition of the Neuroendocrine Stress Response. Journal of Neuroscience. 29:7330–7340.

132. Radley JJ, Johnson SB (2018): Anteroventral bed nuclei of the stria terminalis neurocircuitry: Towards an integration of HPA axis modulation with coping behaviors - Curt Richter Award Paper 2017. Psychoneuroendocrinology. 89:239–249.

133. Vogel SC, Perry RE, Brandes-Aitken A, Braren S, Blair C (2021): Deprivation and threat as developmental mediators in the relation between early life socioeconomic status and executive functioning outcomes in early childhood. Dev Cogn Neurosci. 47:100907.

134. McLaughlin KA, Sheridan MA, Lambert HK (2014): Childhood adversity and neural development: deprivation and threat as distinct dimensions of early experience. Neurosci Biobehav Rev. 47:578–591.

135. Sheridan MA, McLaughlin KA (2014): Dimensions of early experience and neural development: deprivation and threat. Trends Cogn Sci. 18:580–585.

136. Shin LM, Rauch SL, Pitman RK (2006): Amygdala, Medial Prefrontal Cortex, and Hippocampal Function in PTSD. Annals of the New York Academy of Sciences. 1071:67–79.

137. Keding TJ, Herringa RJ (2015): Abnormal Structure of Fear Circuitry in Pediatric Post-Traumatic Stress Disorder. Neuropsychopharmacology. 40:537–545.

138. Herringa R, Phillips M, Almeida J, Insana S, Germain A (2012): Post-traumatic stress symptoms correlate with smaller subgenual cingulate, caudate, and insula volumes in unmedicated combat veterans. Psychiatry Research: Neuroimaging. 203:139–145.

139. Krüger O, Shiozawa T, Kreifelts B, Scheffler K, Ethofer T (2015): Three distinct fiber pathways of the bed nucleus of the stria terminalis to the amygdala and prefrontal cortex. CORTEX. 66:60–68.

140. Murrough JW, Abdallah CG, Anticevic A, Collins KA, Geha P, Averill LA, et al. (2016): Reduced global functional connectivity of the medial prefrontal cortex in major depressive disorder. Hum Brain Mapp. 37:3214–3223.

141. Helpman L, Papini S, Chhetry BT, Shvil E, Rubin M, Sullivan GM, et al. (2016): PTSD REMISSION AFTER PROLONGED EXPOSURE TREATMENT IS ASSOCIATED WITH ANTERIOR CINGULATE CORTEX THINNING AND VOLUME REDUCTION. Depress Anxiety. 33:384–391.

142. Ramirez-Mahaluf JP, Perramon J, Otal B, Villoslada P, Compte A (2018): Subgenual anterior cingulate cortex controls sadness-induced modulations of cognitive and emotional network hubs. Sci Rep. 8:8566.

143. Gianaros PJ, Sheu LK, Matthews KA, Jennings JR, Manuck SB, Hariri AR (2008): Individual differences in stressor-evoked blood pressure reactivity vary with activation, volume, and functional connectivity of the amygdala. Journal of Neuroscience. 28:990.

144. Oler JA, Tromp DPM, Fox AS, Kovner R, Davidson RJ, Alexander AL, et al. (2017): Connectivity between the central nucleus of the amygdala and the bed nucleus of the stria terminalis in the non-human primate: neuronal tract tracing and developmental neuroimaging studies. Brain Structure and Function. 222:21–39.

145. Gray TS, Magnuson DJ (1987): Neuropeptide neuronal efferents from the bed nucleus of the stria terminalis and central amygdaloid nucleus to the dorsal vagal complex in the rat. J Comp Neurol. 262:365–374.

146. Saha S, Batten TFC, Henderson Z (2000): A GABAergic projection from the central nucleus of the amygdala to the nucleus of the solitary tract: a combined anterograde tracing and electron microscopic immunohistochemical study. Neuroscience. 99:613–626.

147. Dong H-W, Swanson LW (2004): Organization of axonal projections from the anterolateral area of the bed nuclei of the stria terminalis. J Comp Neurol. 468:277–298.

148. Maita I, Bazer A, Blackford JU, Samuels BA (2021): Chapter 27 Functional anatomy of the bed nucleus of the stria terminalis–hypothalamus neural circuitry: Implications for valence surveillance, addiction, feeding, and social behaviors. Handb Clin Neurology. 179:403–418.

149. Crestani CC, Alves FHF, Tavares RF, CorrÍa FMA (2009): Role of the bed nucleus of the stria terminalis in the cardiovascular responses to acute restraint stress in rats. Stress: The International Journal on the Biology of Stress. 12:268–278.

150. Brindle RC, Pearson A, Ginty AT (2022): Adverse childhood experiences (ACEs) relate to blunted cardiovascular and cortisol reactivity to acute laboratory stress: A systematic review and meta-analysis. Neurosci Biobehav Rev. 134:104530.

151. Keogh TM, Howard S, Gallagher S (2022): Early Life Adversity and Blunted Cardiovascular Reactivity to Acute Psychological Stress: The Role of Current Depressive Symptoms. Psychosom Med. 84:170–178.

152. Carroll D, Ginty AT, Whittaker AC, Lovallo WR, de Rooij SR (2017): The behavioural, cognitive, and neural corollaries of blunted cardiovascular and cortisol reactions to acute psychological stress. Neurosci Biobehav Rev. 77:74–86.

153. Phillips AC (2011): Blunted cardiovascular reactivity relates to depression, obesity, and self-reported health. Biol Psychol. 86:106–113.

154. Ginty AT, Williams SE, Jones A, Roseboom TJ, Phillips AC, Painter RC, et al. (2016): Diminished heart rate reactivity to acute psychological stress is associated with enhanced carotid intima-media thickness through adverse health behaviors. Psychophysiology. 53:769–775.

155. Gianaros PJ, Kraynak TE, Kuan DCH, Gross JJ, McRae K, Hariri AR, et al. (2020): Affective brain patterns as multivariate neural correlates of cardiovascular disease risk. Social Cognitive and Affective Neuroscience. 15:1034–1045.

