## Supplemental Material for "Amygdala and Subgenual Cingulate Stressor-Evoked Activity Varies Across a Spectrum of Childhood Adversity Severity: Links to Affective and Cardiovascular Outcomes"

3811 O'Hara St

Pittsburgh, PA 15213

**Running title**: Adversity and Central Visceral Circuit Stress Responses

**Keywords**: Childhood Adversity; Amygdala; Subgenual Anterior Cingulate Cortex; Bed Nucleus of the Stria Terminalis; Affect; Cardiovascular

### Methods and Materials

#### Participants

Exclusion criteria included MRI contraindications, pregnancy, left-handedness, cardiovascular disease, diabetes, neurological disorders, use of medications affecting cardiovascular or neural function, suicidality or marked functional impairment and current bipolar, psychotic, or substance use disorders, as previously described(1, 2). Additionally, at the first visit, continued eligibility was determined using medical history, two-week medication history, current substance use and traumatic brain injury inventories.

#### Measures

Table S1. Correlations between Childhood Threat and Deprivation Measures (n=97)

| | | CTQ Threat | THQ 0-11 | THQ 12-17 | CTQ Deprivation | SED |
|---|---|---|---|---|---|---|
| CTQ Threat (Abuse) | Pearson r | -- | | | | |
| | *p* (2-tailed) | | | | | |
| THQ 0-11 (Repeated) | Pearson r | .618 | -- | | | |
| | *p* (2-tailed) | **<.001** | | | | |
| THQ 12-17 (Repeated) | Pearson r | .629 | .735 | -- | | |
| | *p* (2-tailed) | **<.001** | **<.001** | | | |
| CTQ Deprivation (Neglect) | Pearson r | .758 | .423 | .529 | -- | |
| | *p* (2-tailed) | **<.001** | **<.001** | **<.001** | | |
| SED | Pearson r | .385 | .189 | .208 | .376 | -- |
| | *p* (2-tailed) | **<.001** | .064 | **.041** | **<.001** | |

CTQ, Childhood Trauma Questionnaire; SED, Socioeconomic Deprivation; THQ, Trauma History Questionnaire.

### Sample Characterization

Table S2. Summary of Participant Characteristics (n=97)

| Characteristic | Mean | Standard Deviation | Range |
|---|---|---|---|
| Age (years) | 27.32 | 4.02 | 21-35 |
| CTQ Total | 52.43 | 20.86 | 25-100 |
| CTQ Threat (Abuse) | 31.27 | 13.48 | 15-69 |
| CTQ Deprivation (Neglect) | 21.16 | 8.71 | 10-46 |
| THQ (age 0-11, repeated) | .91 | 1.42 | 0-7 |
| THQ (age 12-17, repeated) | 1.13 | 1.85 | 0-11 |
| THQ (>18) | 3.35 | 2.93 | 0-12 |
| Parental education (maximum) | 5.29 | 2.03 | 1-8 |
| Education Level | 5.15 | 1.57 | 1-8 |
| PHQ Depression | 5.19 | 5.50 | 0-22 |
| BDI-II Total | 11.57 | 10.88 | 0-49 |
| PCL-C Total | 32.36 | 14.09 | 17-75 |
| Perceived Stress | 16.88 | 8.85 | 2-37 |
| STAI-T | 41.69 | 13.36 | 20-77 |
| NEO: Neuroticism (%) | 24.73 | 10.16 | 1-47 |
| NEO: Extraversion (%) | 27.34 | 10.16 | 0-44 |
| NEO: Openness (%) | 34.92 | 7.86 | 15-48 |
| NEO: Agreeableness (%) | 32.38 | 7.16 | 12-45 |
| NEO: Conscientiousness (%) | 31.77 | 6.14 | 8-47 |

CTQ, Childhood Trauma Questionnaire; THQ, Trauma History Questionnaire; BDI-II, Beck Depression Inventory-II; NEO, NEO Five-Factor Inventory-3 (NEO-FFI-3); PCL-C, PTSD Checklist-Civilian Version; PHQ, Patient Health Questionnaire; STAI-T, State-Trait Anxiety Inventory Trait score (STAI-Y2).

### fMRI Stressor Task: Multisource Interference Task

During the Multisource Interference Task (MSIT), participants identify the number that differs among three numbers by pressing corresponding positions on a glove. For trials in the congruent (control) condition, the target number in the display appears in a location compatible with its response position. For trials in the incongruent condition, the target number appears in a location incompatible with its response position and accuracy is performance titrated at ~50% by varying the inter-trial intervals (ITI). Thus, greater accuracy within the incongruent blocks prompts shorter ITIs and response time windows; participants were not aware that their performance would be titrated during the MRI protocol. The MSIT is 9 minutes and 20 seconds in duration, comprised of eight alternating incongruent and congruent blocks (four per condition) lasting 52 to 60 seconds. A 10 to 17 second fixation period follows each block. Failure to respond within the designated response time window elicited a "TOO LATE!" and incorrect responses elicited a red "X" as negative feedback, as described previously(3-6). The MSIT resulted in significant anxious arousal, less perceived control and less positive valence in the current sample (Fig. S1, below).

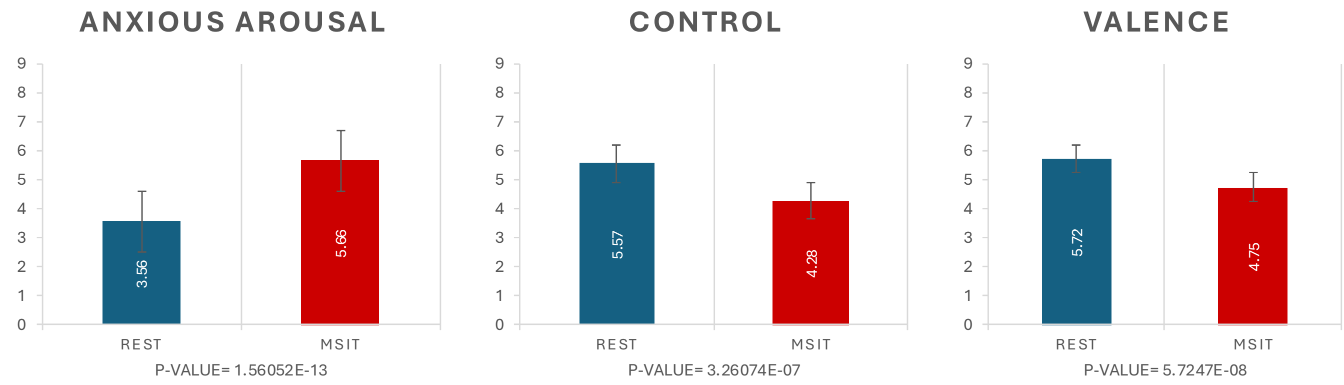


**Figure S1. Subjective reports following rest and the Multisource Interference Task (MSIT)**. Ratings of (A) anxious arousal (1=very calm to 9=very anxious), (B) perceived psychological control (1=very little to 9=very much), and valence (1=very unhappy to 9=very happy) (C) were obtained during the MRI protocol using a 9-point rating scale (Self-Assessment Manikin (SAM)) before and after the Multisource Interference Task. The current sample displayed expected significant differences pre- to post-MSIT (i.e., greater anxious arousal, less perceived control and less positive valence).

### MRI Protocol and Data Acquisition

A custom, localized shimming procedure was implemented that extended from the bottom-most slice to the ventral aspect of the corpus callosum. The FOV was angled 15-20° to ensure visualization of our regions of interest (ROIs). Anatomical images were acquired using a 4.8-min T1-weighted sagittal MPRAGE sequence (TR = 1500 ms, TE = 3.19 ms, flip angle = 8°, 176 slices, FoV = 256 × 256 $mm^2$, voxel size = 1 × 1 × 1.0 $mm^3$). Task-related functional MRI data were acquired using a T2*-weighted gradient-echo echoplanar imaging (EPI) sequence (TR = 2000 ms, TE = 29 ms, flip angle = 65°, slices = 22, Multiband Factor = 3, FoV = 220 × 220 $mm^2$, voxel size = 2 × 2 × 2.0 $mm^3$).

### Preprocessing

A voxel displacement map (VDM) was calculated using a field map image collected immediately prior to the task sequence. Motion correction was applied through realignment of each BOLD image to the first reference image by 6-parameter rigid body transformation. Images were unwarped by applying the previously calculated VDM and resliced. The structural image was then coregistered to the mean functional image. Segmentation was performed on the structural image using probability maps for six tissue classes, generating a deformation field that was then applied to the functional images during normalization of all images to standard MNI space. Smoothing was applied to functional images using a 4 mm full-width-at-half-maximum Gaussian kernel.

**Level 1 Analyses**

Using the block-design task conditions as regressors, task-related activation was modeled by convolving boxcar waveforms with the default SPM canonical hemodynamic response function (HRF). Motion parameters estimated from the realignment preprocessing step were included as additional regressors to account for BOLD signal variance related to head movement. A high-pass filter of 187 seconds was also applied to remove low-frequency BOLD signal noise related to physiological and scanner drift artifact. Model parameters were estimated after accounting for serial correlations using the RobustWLS toolbox, which applied a two-pass variance inverse re-weighting algorithm to further minimize noise artifact(7). After parameter estimation, statistical parametric maps were generated for specified T-contrasts comparing relative BOLD activation between the congruent, incongruent and fixation conditions.

**Level 2 Analysis**

To examine task-related activity, a random effects analysis was performed using a one-sample t-test. First-level contrast maps were averaged across all subjects to generate an averaged contrast map for the incongruent vs. congruent condition. After model parameter estimation, second-level T-contrasts were specified using a positive-weighted vector [1] and a negative-weighted vector of [-1] to examine both significant stressor-evoked activation and deactivation. Cluster maps showing significant changes in activity across the whole brain were generated from the resulting statistical parametric maps using a false discovery rate (FDR) significance threshold of 0.05 to correct for multiple comparisons, and a cluster extent threshold of 20 voxels.

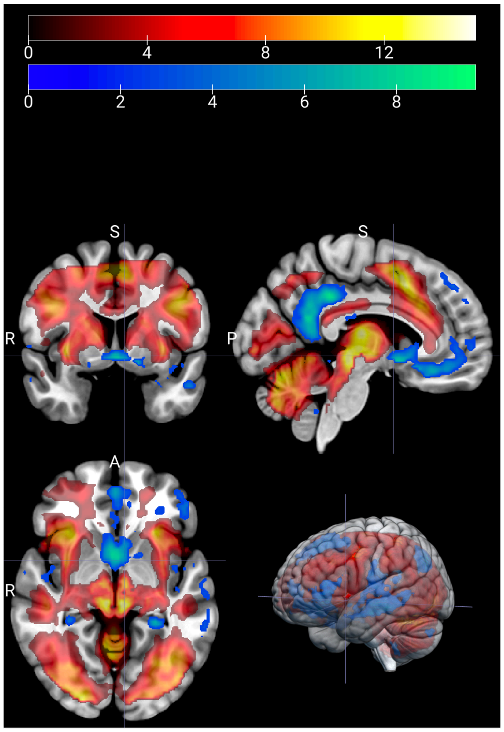


**Figure S2. Multisource Interference Task (MSIT) Main Effects Maps.** Significant whole-brain task-related deactivation (blue-green) and activation (red-yellow) displayed on coronal (top, left), sagittal (top, right), and axial sections (bottom, left), along with surface rendering (bottom, right).

#### Region of Interest Analyses

Clusters were identified at an extent threshold of 10 voxels for the amygdala, BNST and sgACC, and 0 voxels for the PVN, due to its small ROI size. The BOLD signal change within each ROI for each subject was computed by extracting the first eigenvariate from each cluster. Significant deactivation (lower BOLD signal in the incongruent vs. congruent condition) was detected in all ROIs: BNST (left, right k=14, 12), PVN (k=2, 2), sgACC (k=79), and amygdala (k=32, 25). Significant activation (higher BOLD signal in the incongruent vs. congruent condition) was detected only in the amygdala (k=28, 43). Overlap between whole-brain deactivation and ROIs has been identified (Fig. S3).

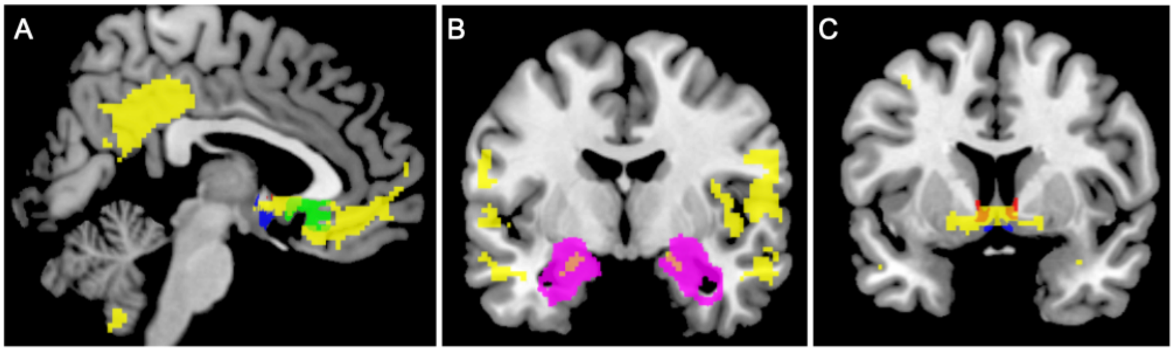


**Figure S3. Multisource Interference Task (MSIT) Whole-Brain Activity and Overlap with Regions of Interest (ROIs).** Significant whole-brain deactivation (yellow) overlying ROIs: A (sagittal). sgACC (green, sagittal), B. (coronal) amygdala (magenta), C. (coronal) bed nucleus of the stria terminalis (red), and paraventricular nucleus of the hypothalamus (blue).

**Stressor-Evoked Connectivity: Generalized Psychophysiological Interaction Analyses (gPPI)**

To measure significant task-dependent connectivity between our four ROIs, a volume of interest (VOI) was generated for each ROI for each subject. The first eigenvariate of the BOLD signal time series was generated and extracted from each VOI using singular value decomposition from fMRI data during the task. Using the SPM8 Generalized PPI Toolbox(8), functional connectivity analyses were estimated from each ROI seed to the rest of the brain. The principal time series of each ROI seed region, task condition (fixation, congruent or incongruent) convolved with the SPM8 hemodynamic response function (HRF), and motion parameters were entered as regressors in each individual GLM design matrix. Individual GLMs were estimated, generating PPI contrast maps for the main task condition (incongruent vs. congruent). Individual GLM testing demonstrates time-dependent co-variation (i.e. functional connectivity) between the seed region and the rest of the brain for each subject.

Using the SnPM Toolbox statistical nonparametric permutation-based mapping method (for SPM12; http://nisox.org/Software/SnPM13/), independent samples t-tests were performed averaging the second-level PPI contrasts to determine significantly functionally connected voxels across subjects within each ROI. We used 5,000 permutations to calculate non-parametric p-values with a cluster-forming threshold of $p<0.001$ to control for multiple comparisons at a false discovery rate (FDR) corrected voxel-level threshold of $<0.05$. An activation map was then generated for each ROI seed and masked with each of the other ROIs, generating a cluster of significant task-related PPI effect between regions. The first eigenvariate of the PPI was extracted from each activation cluster map to examine the relationship between childhood adversity measures and stressor-evoked functional connectivity. *Negative* connectivity values indicate *greater* task-related coupling during incongruent compared to congruent conditions, while *positive* values indicate *greater* task-related coupling during congruent compared to incongruent conditions, and near-zero connectivity reflects minimal differential coupling between conditions.

Applying an activation cluster voxel threshold of k=10, in the congruent condition, significant functional connectivity was found between the following regions [seed-mask]: amygdala-BNST (left, right k=21, 21), BNST-amygdala (k=137, 232), BNST-sgACC (k=39), PVN-amygdala (k=80, 134), PVN-BNST (k=17, 18), sgACC-amygdala (k=65, 140), sgACC-BNST (k=15, 16).

### Diagnostic Assessment

Of the 69 participants with a history of affective diagnosis, 46 participants had one or more current affective diagnoses. Prevalent primary lifetime diagnoses were trauma-related disorder (n=29), depressive disorder (n=24) and anxiety disorder (n=16). Further, 36 had comorbid lifetime mood and anxiety/trauma-related disorders.

### Results

### Childhood Adversity and Affective or Cardiovascular Outcomes

Greater childhood adversity was significantly correlated with greater affective symptoms and disorders. CTQ Threat showed large associations, THQ 0-11 showed small-to-moderate associations, and socioeconomic deprivation showed moderate associations (Table S3). Greater childhood threat was significantly correlated with greater SBP reactivity (larger blood pressure increases from baseline to stress) and impaired DBP recovery (smaller decreases in DBP from stress to recovery) with small to moderate effect sizes (Table S4).

Table S3. Childhood Adversity and Affective Outcomes

| | | CTQ Threat | THQ 0-11 | SED | BDI-II Total | PCL Total | Lifetime Diagnoses |
|---|---|---|---|---|---|---|---|
| CTQ Threat | Pearson r | -- | | | | | |
| | *p* (2-tailed) | | | | | | |
| THQ 0-11 (Repeated) | Pearson r | .618 | | | | | |
| | *p* (2-tailed) | **<.001** | | | | | |
| SED | Pearson r | .385 | .189 | -- | | | |
| | *p* (2-tailed) | **<.001** | .064 | | | | |
| BDI-II Total | Pearson r | .529 | .282 | .347 | -- | | |
| | *p* (2-tailed) | **<.001** | **.005** | **<.001** | | | |
| PCL Total | Pearson r | .562 | .411 | .296 | .851 | -- | |
| | *p* (2-tailed) | **<.001** | **<.001** | **.003** | **<.001** | | |
| Lifetime Diagnoses | Pearson r | .629 | .426 | .363 | .689 | .702 | -- |
| | *p* (2-tailed) | **<.001** | **<.001** | **<.001** | **<.001** | **<.001** | |

BDI-II, Beck Depression Inventory-II; CTQ, Childhood Trauma Questionnaire; PCL-C, PTSD Checklist-Civilian Version; SED, Socioeconomic Deprivation; THQ, Trauma History Questionnaire.

Table S4. Childhood Adversity and Cardiovascular Outcomes

| | | CTQ Threat | THQ 0-11 | SED | Baseline SBP | SBP Reactivity | SBP Rec | Baseline DBP | DBP Reactivity | DBP Rec | Baseline HR | HR Reactivity | HR Rec |
|---|---|---|---|---|---|---|---|---|---|---|---|---|---|
| CTQ Threat | Pearson r | -- | | | | | | | | | | | |
| | *p* (2-tailed) | | | | | | | | | | | | |
| THQ 0-11 | Pearson r | .618 | -- | | | | | | | | | | |
| | *p* (2-tailed) | **<.001** | | | | | | | | | | | |
| SED | Pearson r | .385 | .189 | -- | | | | | | | | | |
| | *p* (2-tailed) | **<.001** | .064 | | | | | | | | | | |
| Baseline SBP | Pearson r | .023 | -.161 | .047 | -- | | | | | | | | |
| | *p* (2-tailed) | .824 | .124 | .655 | | | | | | | | | |
| SBP Reactivity | Pearson r | .025 | .237 | .081 | -.101 | -- | | | | | | | |
| | *p* (2-tailed) | .812 | **.023** | .444 | .337 | | | | | | | | |
| SBP Recovery | Pearson r | .182 | .021 | -.029 | -.270 | -.416 | -- | | | | | | |
| | *p* (2-tailed) | .081 | .843 | .780 | **.010** | **<.001** | | | | | | | |
| Baseline DBP | Pearson r | -.026 | -.148 | .034 | .895 | -.057 | -.190 | -- | | | | | |
| | *p* (2-tailed) | .805 | .157 | .745 | **<.001** | .590 | .071 | | | | | | |
| DBP Reactivity | Pearson r | -.016 | .086 | .045 | -.220 | .594 | -.111 | -.237 | -- | | | | |
| | *p* (2-tailed) | .883 | .415 | .667 | **.035** | **<.001** | .295 | **.023** | | | | | |
| DBP Recovery | Pearson r | .360 | .172 | .077 | -.051 | -.204 | .645 | .013 | -.317 | -- | | | |
| | *p* (2-tailed) | **<.001** | .100 | .466 | .629 | .053 | **<.001** | .906 | **.002** | | | | |
| Baseline HR | Pearson r | .088 | -.097 | -.013 | .165 | -.009 | -.085 | .180 | -.218 | .105 | -- | | |
| | *p* (2-tailed) | .403 | .356 | .902 | .114 | .932 | .425 | .084 | **.037** | .323 | | | |
| HR Reactivity | Pearson r | .022 | .064 | -.034 | -.172 | .390 | -.160 | -.149 | .200 | -.156 | -.103 | -- | |
| | *p* (2-tailed) | .837 | .545 | .750 | .100 | **<.001** | .130 | .156 | .056 | .140 | .329 | | |
| HR Recovery | Pearson r | .011 | -.130 | .010 | -.066 | -.341 | .424 | -.124 | -.169 | .275 | -.329 | -.391 | -- |
| | *p* (2-tailed) | .918 | .216 | .927 | .535 | **<.001** | **<.001** | .240 | .110 | **.008** | **.001** | **<.001** | |

CTQ, Childhood Trauma Questionnaire; DBP, Diastolic Blood Pressure; HR, Heart Rate; Rec, Recovery; SBP, Systolic Blood Pressure; SED, Socioeconomic Deprivation; THQ, Trauma History Questionnaire.

**Childhood Adversity and CVC Stressor-Evoked Activity/Connectivity**

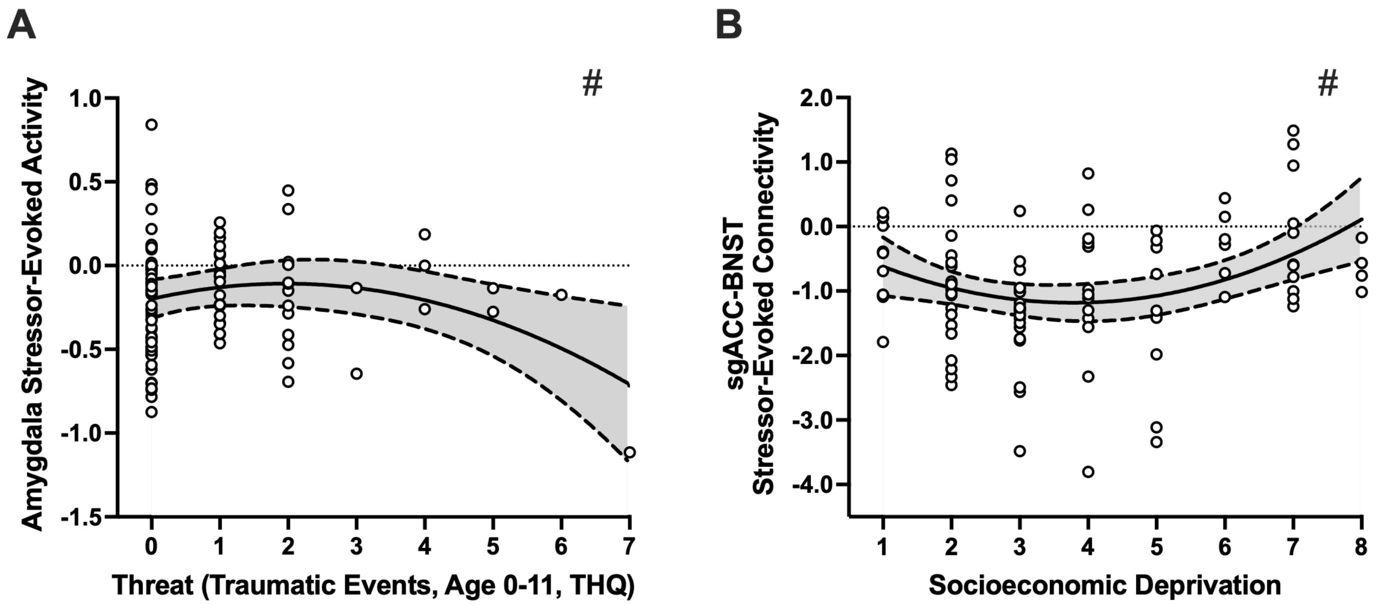


**Figure S4. Childhood adversity and Central Visceral Circuit stressor-evoked activity and connectivity.** **(A)** THQ 0–11 showed a significant inverted U-shaped relationship with amygdala activity, with amygdala reactivity relatively blunted across low-to-moderate trauma exposure, increasing at the highest levels of exposure (i.e., greater deactivation; β=-0.546, *P*=0.033). **(B)** SED showed a significant U-shaped relationship with sgACC-BNST stressor-evoked connectivity when evaluated in a curvilinear model (CTQ Threat model: β=1.067, *P*=0.028; THQ 0-11 model: β=1.071, *P*=0.022), with weaker (near-zero) connectivity at low and high deprivation and greater connectivity at moderate deprivation. #Indicates the association was significant in the full model with the adulthood covariates (traumatic events >18 years, education level/SES, past-year negative life events). (BNST, bed nucleus of the stria terminalis; sgACC, subgenual anterior cingulate cortex; THQ, Trauma History Questionnaire).

**Childhood Threat and Amygdala Stressor-Evoked Activity: Examination of Potential Outliers or Points of Leverage**

Residual plots identified one potentially high-leverage observation (THQ 0-11 = 7, highest observed value) with elevated standardized residual. The influence of this case differed between threat models: in the THQ 0-11 model, this participant was isolated at the extreme of the distribution, whereas in the CTQ model, the same participant appeared closer to other observations, reducing their relative influence. Using the outlier labeling method, we determined that there are *no* outliers based on the dependent variable (i.e., stressor-evoked amygdala activity). Sensitivity analyses excluding this participant demonstrate that the finding is maintained in the CTQ Threat model but not in the THQ 0-11 model (Table S5).

Given the positive skew of THQ 0-11 scores (skewness = 2.14, kurtosis = 4.98), we conducted an exploratory generalized linear model with Huber-White robust standard errors(9) to verify the robustness of initial findings. This approach adjusts standard errors to account for distributional properties and potential influential cases while maintaining the original scale of measurement. The robust model showed excellent fit (Deviance/df = 0.108) and yielded similar results to those of the reported regression. The quadratic effect of THQ 0-11 remained significant (THQ $0\text{-}11^2$: β=-0.025, SE=0.012, *P*=0.031) in the full model, demonstrating that the curvilinear relationship is not driven by extreme values.

Table S5. Results of Sensitivity Analyses without THQ 0-11 = 7 (n=96)

| Model | Step | Variable | St. β | t | *p* |
|---|---|---|---|---|---|
| CTQ Threat vs. Amygdala Activity (Fig. 1A) | 3 | **CTQ Threat²** | -1.211 | -2.314 | **.023** |
| | 4 | **CTQ Threat²** | -1.394 | -2.506 | **.014** |
| THQ 0-11 vs. Amygdala Activity (Fig. S4) | 3 | **THQ 0-11²** | -.150 | -.568 | .572 |
| | 4 | **THQ 0-11²** | -.169 | -.624 | .534 |

CTQ, Childhood Trauma Questionnaire; THQ, Trauma History Questionnaire.

### CVC Stressor-Evoked Activity/Connectivity and Affective Symptoms and Disorders

Table S6. Regression Results: Amygdala Stressor-Evoked Activity and Post-Traumatic Stress Symptoms

| | | **Post-traumatic Stress Symptoms (PCL-C)** | | |
|---|---|---|---|---|
| Step | Variable | St. β | t | *p* |
| 1 | Age | .193 | 1.880 | .063 |
| | Sex | -.134 | -1.302 | .196 |
| | Race | .020 | .190 | .850 |
| | Amygdala Stressor-Evoked Activity | .067 | .646 | .520 |
| 2 | Age | .183 | 1.791 | .077 |
| | Sex | -.158 | -1.529 | .130 |
| | Race | .030 | .298 | .766 |
| | Amygdala Stressor-Evoked Activity | -.045 | -.370 | .712 |
| | **Amygdala Stressor-Evoked Activity²** | -.204 | -1.661 | .100 |
| 3 | Age | -.066 | -.609 | .544 |
| | Sex | -.023 | -.237 | .813 |
| | Race | .009 | .099 | .922 |
| | Amygdala Stressor-Evoked Activity | -.015 | -.134 | .894 |
| | **Amygdala Stressor-Evoked Activity²** | -.228 | -2.066 | **.042** |
| | THQ >18 | .280 | 2.453 | **.016** |
| | Adulthood SES | .061 | .654 | .515 |
| | Negative Life Events | .352 | 3.273 | **.002** |

Bold values indicate significance at p<0.05; findings did not survive FDR correction (0.05) for 3 tests. PCL-C, PTSD Checklist-Civilian Version; SES, Socioeconomic Status; THQ, Trauma History Questionnaire.

Table S7. Regression Results: Subgenual ACC Stressor-Evoked Activity and Post-Traumatic Stress Symptoms

| | | **Post-traumatic Stress Symptoms (PCL-C)** | | |
|---|---|---|---|---|
| Step | Variable | St. β | t | *p* |
| 1 | Age | .190 | 1.859 | .066 |
| | Sex | -.126 | -1.236 | .219 |
| | Race | .028 | .275 | .784 |
| 2 | Age | .181 | 1.780 | .078 |
| | Sex | -.131 | -1.288 | .201 |
| | Race | .017 | .171 | .864 |
| | **sgACC Stressor-Evoked Activity** | .150 | 1.489 | .140 |
| 3 | Age | -.068 | -.626 | .533 |
| | Sex | .008 | .079 | .937 |
| | Race | -.001 | -.010 | .992 |
| | **sgACC Stressor-Evoked Activity** | .184 | 1.988 | **.050** |
| | THQ >18 | .281 | 2.471 | **.015** |
| | Adulthood SES | .070 | .743 | .459 |
| | Negative Life Events | .344 | 3.191 | **.002** |

Bold values indicate significance at p<0.05; findings did not survive FDR correction (0.05) for 3 tests. sgACC, subgenual anterior cingulate cortex; PCL-C, PTSD Checklist-Civilian Version; SES, Socioeconomic Status; THQ, Trauma History Questionnaire.

Table S8. Regression Results: BNST-sgACC Stressor-Evoked Connectivity and Lifetime Diagnoses

| | | **Lifetime Diagnoses** | | |
|---|---|---|---|---|
| Step | Variable | St. β | t | *p* |
| 1 | Age | .335 | 3.457 | **.001** |
| | Sex | -.290 | -3.041 | **.003** |
| | Race | -.039 | -.410 | .683 |
| | BNST-sgACC Stressor-Evoked Connectivity | .036 | .372 | .711 |
| 2 | Age | .339 | 3.564 | **.001** |
| | Sex | -.302 | -3.224 | **.002** |
| | Race | -.061 | -.644 | .521 |
| | BNST-sgACC Stressor-Evoked Connectivity | -.164 | -1.238 | .219 |
| | **BNST-sgACC Stressor-Evoked Connectivity$^2$** | -.285 | -2.145 | **.035** |
| 3 | Age | .130 | 1.260 | .211 |
| | Sex | -.189 | -2.056 | **.043** |
| | Race | -.090 | -1.024 | .309 |
| | BNST-sgACC Stressor-Evoked Connectivity | -.190 | -1.544 | .126 |
| | **BNST-sgACC Stressor-Evoked Connectivity$^2$** | -.280 | -2.281 | **.025** |
| | THQ >18 | .250 | 2.325 | **.022** |
| | Adulthood SES | -.075 | -.848 | .399 |
| | Negative Life Events | .243 | 2.387 | **.019** |

Bold values indicate significance at p<0.05; findings did not survive FDR correction (0.05) for 3 tests. BNST, bed nucleus of the stria terminalis; sgACC, subgenual anterior cingulate cortex; SES, Socioeconomic Status; THQ, Trauma History Questionnaire.

### CVC Stressor-Evoked Activity and Cardiovascular Outcomes

Table S9. Regression Results: Amygdala Stressor-Evoked Activity and DBP Reactivity and Recovery

| | | **DBP Reactivity** | | | **DBP Recovery** | | |
|---|---|---|---|---|---|---|---|
| Step | Variable | St. β | t | *p* | St. β | t | *p* |
| 1 | Age | .026 | .239 | .812 | .006 | .055 | .956 |
| | Sex | .066 | .612 | .542 | -.290 | -2.867 | **.005** |
| | Race | -.011 | -.102 | .919 | .120 | 1.184 | .240 |
| 2 | Age | .038 | .361 | .719 | -.007 | -.067 | .947 |
| | Sex | .032 | .307 | .759 | -.256 | -2.608 | **.011** |
| | Race | -.046 | -.436 | .664 | .155 | 1.577 | .118 |
| | **Amygdala Stressor-Evoked Activity** | .272 | 2.604 | **.011*** | -.277 | -2.815 | **.006*** |
| 3 | Age | .020 | .164 | .870 | .041 | .364 | .717 |
| | Sex | .007 | .060 | .952 | -.217 | -2.107 | **.038** |
| | Race | -.040 | -.372 | .711 | .146 | 1.499 | .137 |
| | **Amygdala Stressor-Evoked Activity** | .286 | 2.678 | **.009*** | -.304 | -3.105 | **.003*** |
| | THQ >18 | .082 | .627 | .532 | -.174 | -1.445 | .152 |
| | Adulthood SES | .099 | .924 | .358 | -.179 | -1.818 | .073 |
| | Negative Life Events | -.074 | -.603 | .548 | .122 | 1.082 | .282 |

Bold values indicate significance at p<0.05; an asterisk indicates survival of FDR correction (0.05) for 3 tests. DBP, Diastolic Blood Pressure; SES, Socioeconomic Status; THQ, Trauma History Questionnaire.

Table S10. Regression Results: Amygdala-BNST Stressor-Evoked Connectivity and SBP Reactivity

| | | SBP Reactivity | | |
|---|---|---|---|---|
| Step | Variable | St. β | t | *p* |
| 1 | Age | .075 | .694 | .490 |
| | Sex | .035 | .323 | .747 |
| | Race | .034 | .320 | .749 |
| | Amygdala-BNST Stressor-Evoked Connectivity | -.016 | -.149 | .882 |
| 2 | Age | .085 | .800 | .426 |
| | Sex | .013 | .120 | .905 |
| | Race | -.022 | -.203 | .840 |
| | Amygdala-BNST Stressor-Evoked Connectivity | -.408 | -1.999 | **.049** |
| | **Amygdala-BNST Stressor-Evoked Connectivity$^2$** | -.458 | -2.242 | **.028** |
| 3 | Age | .036 | .291 | .772 |
| | Sex | .049 | .434 | .666 |
| | Race | -.010 | -.093 | .926 |
| | Amygdala-BNST Stressor-Evoked Connectivity | -.445 | -2.095 | **.039** |
| | **Amygdala-BNST Stressor-Evoked Connectivity$^2$** | -.514 | -2.423 | **.018** |
| | THQ >18 | .025 | .190 | .850 |
| | Adulthood SES | .164 | 1.509 | .135 |
| | Negative Life Events | .140 | 1.117 | .267 |

Bold values indicate significance at p<0.05; findings did not survive FDR correction (0.05) for 3 tests. BNST, bed nucleus of the stria terminalis; SBP, Systolic Blood Pressure; SES, Socioeconomic Status; THQ, Trauma History Questionnaire.

### Amygdala-BNST Connectivity and SBP Reactivity: Examination of Potential Outliers or Points of Leverage

Residual plots identified an observation at the extreme of the predictor space showing an elevated standardized residual; this participant has an Amygdala-BNST connectivity = -4.510 (the lowest value). Using the outlier labeling method, we determined that this point is a marginal outlier (Lower-Upper bounds: -4.355 - 1.956). We have included sensitivity analyses below; findings become null when excluding this individual but remain significant when Winsorizing to the next lowest value. Although not a point of leverage, there was also a marginal outlier for SBP reactivity = 16.667 (Lower-Upper bounds: -11.075 - 16.2625). Sensitivity analyses excluding and Winsorizing this point are below (Table S11).

To further evaluate robustness, we employed an exploratory generalized linear model with Huber-White robust standard errors, which adjust for potential influence of extreme values while retaining all observations. The robust model yielded similar coefficients to the reported results in the full model (Amygdala-BNST$^2$: β=-.603, SE=0.296, *P*=0.042), demonstrating that the curvilinear relationship between amygdala-BNST connectivity and SBP reactivity is not overly driven by extreme values.

Table S11. Sensitivity Analyses for Amygdala-BNST Connectivity and SBP Reactivity

| Model | Outlier | Outlier Method | Step | Variable | St. β | t | *p* |
|---|---|---|---|---|---|---|---|
| Amygdala-BNST Connectivity vs. SBP Reactivity (Fig. 3C) | Amygdala-BNST Outlier (-4.51) | Exclusion | 2 | **Amygdala-BNST Connectivity[2]** | -.104 | -.506 | .614 |
| | | | 3 | | -.165 | -.763 | .448 |
| | | Winsorization to the next value (-4.25) | 2 | | -.465 | -2.331 | **.022** |
| | | | 3 | | -.518 | -2.507 | **.014** |
| | SBP Reactivity outlier (16.67) | Exclusion | 2 | | -.467 | -2.265 | **.026** |
| | | | 3 | | -.522 | -2.458 | **.016** |
| | | Winsorization to the next value (16.25) | 2 | | -.459 | -2.248 | **.027** |
| | | | 3 | | -.515 | -2.430 | **.017** |
| | Both Outliers | Exclusion | 2 | | -.086 | -.419 | .677 |
| | | | 3 | | -.148 | -.687 | .494 |
| | | Winsorization | 2 | | -.466 | -2.337 | **.022** |
| | | | 3 | | -.519 | -2.514 | **.014** |

BNST, bed nucleus of the stria terminalis; SBP, Systolic Blood Pressure.

### References


1. Banihashemi L, Peng CW, Rangarajan A, Karim HT, Wallace ML, Sibbach BM, et al. (2022): Childhood Threat Is Associated With Lower Resting-State Connectivity Within a Central Visceral Network. *Front Psychol*. 13:805049.
2. Banihashemi L, Peng CW, Verstynen T, Wallace ML, Lamont DN, Alkhars HM, et al. (2021): Opposing relationships of childhood threat and deprivation with stria terminalis white matter. *Human Brain Mapping*. 42:2445-2460.
3. Gianaros PJ, Sheu LK, Remo AM, Christie IC, Critchley HD, Wang J (2009): Heightened resting neural activity predicts exaggerated stressor-evoked blood pressure reactivity. *Hypertension*. 53:819-825.
4. Gianaros PJ, Onyewuenyi IC, Sheu LK, Christie IC, Critchley HD (2011): Brain systems for baroreflex suppression during stress in humans. *Human Brain Mapping*. 33:1700-1716.
5. Sheu LK, Jennings JR, Gianaros PJ (2012): Test-retest reliability of an fMRI paradigm for studies of cardiovascular reactivity. *Psychophysiology*. 49:873-884.
6. Banihashemi L, Sheu LK, Midei AJ, Gianaros PJ (2015): Childhood physical abuse predicts stressor-evoked activity within central visceral control regions. *Soc Cogn Affect Neurosci*. 10:474-485.
7. Diedrichsen J, Shadmehr R (2005): Detecting and adjusting for artifacts in fMRI time series data. *NeuroImage*. 27:624-634.
8. McLaren DG, Ries ML, Xu G, Johnson SC (2012): A generalized form of context-dependent psychophysiological interactions (gPPI): A comparison to standard approaches. *NeuroImage*. 61:1277-1286.
9. Huber PJ (1967): The behavior of maximum likelihood estimates under nonstandard conditions. *Proceedings of the fifth Berkeley symposium on mathematical statistics and probability*, 1 ed. Berkeley, CA: University of California Press, pp 221-233.
